# Development of AWaRe quality indicators for optimal antibiotic use

**DOI:** 10.1101/2025.10.24.25338539

**Authors:** Annie Heath, Jan Goelen, Pem Chuki, Aislinn Cook, Filip Djukic, Nga Thi Thuy Do, Elisa Funiciello, Sumanth Gaundra, Brian Godman, Benedikt Huttner, Yara Mohsen Khalaf, Guilia Lorenzetti, Marc Mendelson, Catrin E Moore, Claudia GS Osorio-de-Castro, Zikria Saleem, Jeroen Schouten, Elizabeth Tayler, Evelyn Wesangula, AWaRe QIs Delphi Panel, Stephen M Campbell, Michael Sharland

## Abstract

**Background:** The WHO AWaRe (Access/Watch/Reserve) book gives detailed guidance on the optimal use of antibiotics across primary care and hospital settings aiming to improve the quality of antibiotic use.

**Objectives:** To develop model sets of appropriate and feasible quality indicators (QIs) based on the WHO AWaRe system, that are widely implementable across primary care, hospital, and general healthcare system settings to support optimal antibiotic use.

**Methods:** Indicators from a scoping review were revised to focus on clinical infections in the AWaRe book. They were assessed using two rounds each of the Global Delphi Technique and RAND/UCLA Appropriateness Method evaluating appropriateness and feasibility at national and global levels respectively. In Round 1 of each method, panellists rated clarity and suggested revisions or new indicators. Round 2 results are reported. The indicators were then reviewed to create a priority list based on WHO book antibiotic choice recommendations, burden of disease, and positive measurement.

**Results:** The AWaRe QIs covered common primary and hospital infections listed in the AWaRe book and general patient and population level indicators. There were 102 quality indicators (Primary Care: 46; Hospital: 39; General: 17) included in Round 2 of the Delphi Technique and 136 indicators (Primary Care: 56; Hospital: 60; General: 20) in Round 2 of the RAND/UCLA method.. The prioritisation process resulted in 20 core indicators (Primary Care: 10, Hospital: 8, General: 2).

**Conclusion:** AWaRe quality indicators have been derived to support optimal antibiotic use, and inform stewardship programs.

## Introduction

The AWaRe (Access, Watch, Reserve) system was developed by the WHO and includes the classification of antibiotics based on their spectrum of activity and resistance potential. ^1^ The AWaRe system (Figure 1) is aligned with and complements the WHO Model List of Essential Medicines (EML), a tool developed by the WHO that prioritises the use of specific antibiotics. ^2^ It facilitates and promotes appropriate antibiotic use ^3,4^ and underpins the recent United Nations General Assembly (UNGA) target that at least 70% of all human use of antibiotics globally should be Access antibiotics. ^5^ The 2022 AWaRe antibiotic book provides empiric treatment guidance for 34 common infections in primary and hospital care for children and adults based on the AWaRe system. ^6,7^ The AWaRe book provided detailed evidence-based guidance on the choice of drug, dose and duration, and is more comprehensive than antibiotic guidelines from many national guidelines.^8^ Improving appropriate antibiotic use requires insight into current patterns of use at regional, national, and healthcare facility levels through measurement and comparison to agreed standards through validated quality indicators (QI).^9^

**Figure 1:**
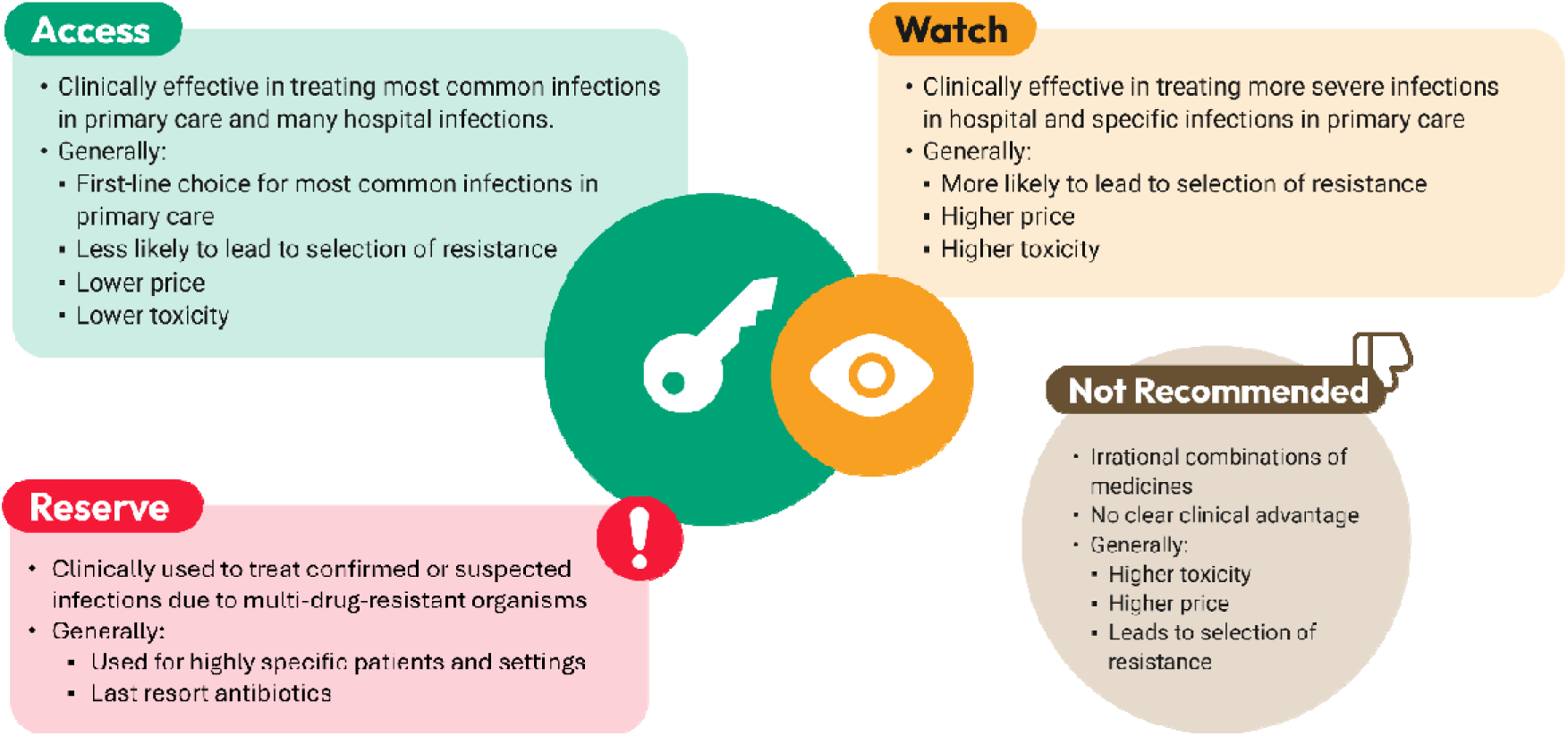
Overview of the WHO AWaRe Groups^7^.

QIs are clearly defined, quantifiable scores based on evidence, which should be tracked over time and demonstrate that greater adherence is associated with better quality of care. ^10,11^ The WHO and other organisations have developed antibiotic stewardship toolkits for hospitals and ambulatory care to improve antibiotic use. ^12,13^ However, there are currently no agreed lists of QIs reflecting recommendations in WHO AWaRe book guidance to assess and optimise the appropriate use of antibiotics. This is especially important for the primary care sector, where over 90% of antibiotics are dispensed in most countries. ^14,15^ As part of the Antibiotic Data to Inform Local Action (ADILA) project (Wellcome Trust grant number: 222051/Z/20/Z), we conducted an initial scoping review that identified 773 existing quality indicators and treatment recommendations on systemic antibiotic use. ^16^ The review found that for infection-specific indicators, approximately 50% focused on respiratory tract infections and there were no indicators for many clinically important infections. The review highlighted that only 8 of 773 (1%) indicators were based on the AWaRe system directly although 445 (57.6%) indicators indirectly referred to the guidance provided in the AWaRe antibiotic book. ^16^ The review demonstrated the need to develop globally applicable indicators linked to the AWaRe book to inform antimicrobial stewardship (AMS) in both primary and hospital settings, for adults and children, with a focus on lower-resource settings.

As part of the ADILA project, this study aimed to develop model sets of appropriate and feasible QIs, based on the WHO AWaRe book guidelines for common infections in primary and hospital care, and general QIs for appropriate antibiotic use, that can be adapted locally as part of AMS activities within individual countries or settings.

## Methods

### Delphi and RAND/UCLA Appropriateness Techniques

The indicators identified in the initial scope ^16^ were reviewed to focus specifically on the clinical infections included in the 2022 WHO AWaRe antibiotic book on empiric antibiotic treatment and management of common infections. ^6^ The general QIs were based on the antibiotic prescribing and dispensing guidance given in the AWaRe antibiotic book on the appropriate use of antibiotics across all settings.

Two distinct consensus methodology processes were conducted, each with a different focus and panel composition.

The first was a two-round Delphi Technique which engaged a panel of over 100 international experts in AMR and AMS from all WHO regions (Box S1). ^17^ This process assessed the appropriateness and feasibility of the indicators within national and local health system contexts (Figure 2).

**Figure 2:**
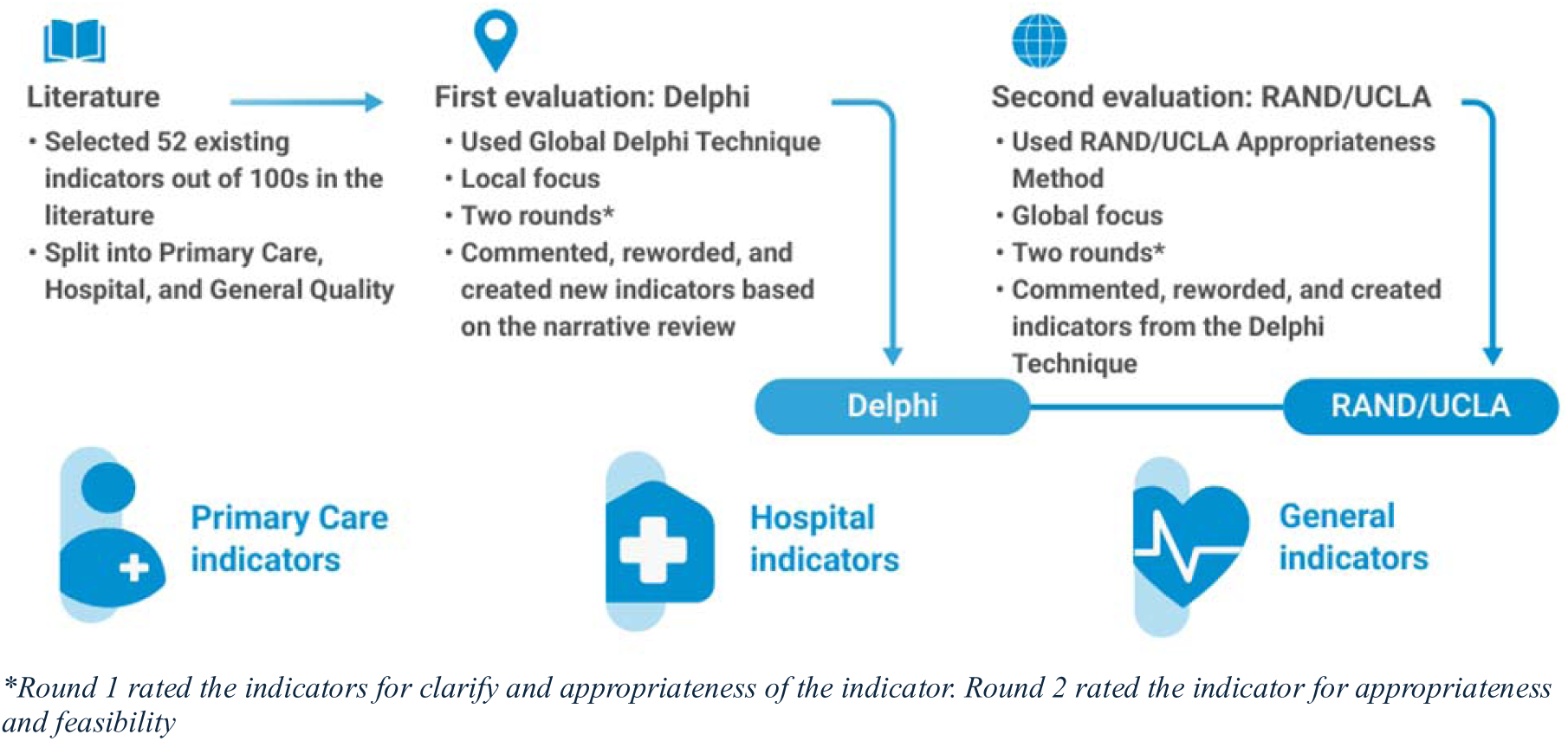
Delphi Technique and RAND/UCLA Method Processes.

Following this, a separate RAND/UCLA Appropriateness Method was conducted with a panel of 12 global experts in AMR/AMS from all WHO regions (PC, NTTD, SG, BH, YMK, FCL, MM, CGSOC, ZS, JS, ET, EW) (Box S1). ^11,18^ This second process focused specifically on evaluating the appropriateness and feasibility of the indicators in a global context, considering their potential for broader international application and comparability (Figure 2).^19^

In Round 1 of each method, the panellists rated indicators on separate 9-point integer scales for the clarity and appropriateness of each indicator, and appropriateness and current feasibility in Round 2 of each method (Figure S1 and Box S2). During the first round, panellists were encouraged to provide comments to improve the wording of the indicators. Any newly worded indicators were included in the second round for rating, in addition to the originally worded indicators. Revised indicators that more clearly captured the intended concept were brought into the RAND/UCLA method for rating. This ensured that feedback from the Delphi panel was reflected in more precise and refined versions in the RAND/UCLA method.

Both rounds of the Delphi Technique and Round 1 of the RAND/UCLA method were conducted online with no interaction between panellists. In Round 2 of the RAND/UCLA method, the panellists met virtually for a 4-hour face-to-face interactive meeting to discuss, and in some cases reword, the indicators for greater clarity and to rate each indicator for appropriateness and feasibility. Any newly worded indicators during the RAND/UCLA meeting were rated anonymously immediately during the meeting using Mentimeter. All other indicators were rated individually on the panellists’ ratings MS Excel® file during the meeting. Detailed outline of the Delphi Technique and RAND/UCLA method can be found in the Supplementary materials (pg. 4).

After the indicators were rated by the panellists, the QIs were then reviewed to first remove duplicates and edit the wording for consistency. Then a priority list was developed that directly reflected the WHO recommendation of choice of antibiotic for specific infections with supporting indicators covering the dose and duration of the recommended antibiotic for that infection in the AWaRe book. Priority QIs also included those that were positive indicators, indicators that measure treatment strategies in line with the AWaRe book and for infections with higher disease burden.^7,20^ All developed indicators and all ratings from the Delphi and RAND/UCLA Round 2 processes are presented in the supplement (pg. 10).

## Results

### Delphi and RAND/UCLA Appropriateness Techniques

Building on the QIs brought forward from the scoping review, ^16^ 52 indicators were rated during Delphi Round 1. An additional 83 QIs were suggested by panellists and were added to the list in Round 2 (Figure 3). This resulted in 102 indicators at the end of the Delphi Round 2.

**Figure 3:**
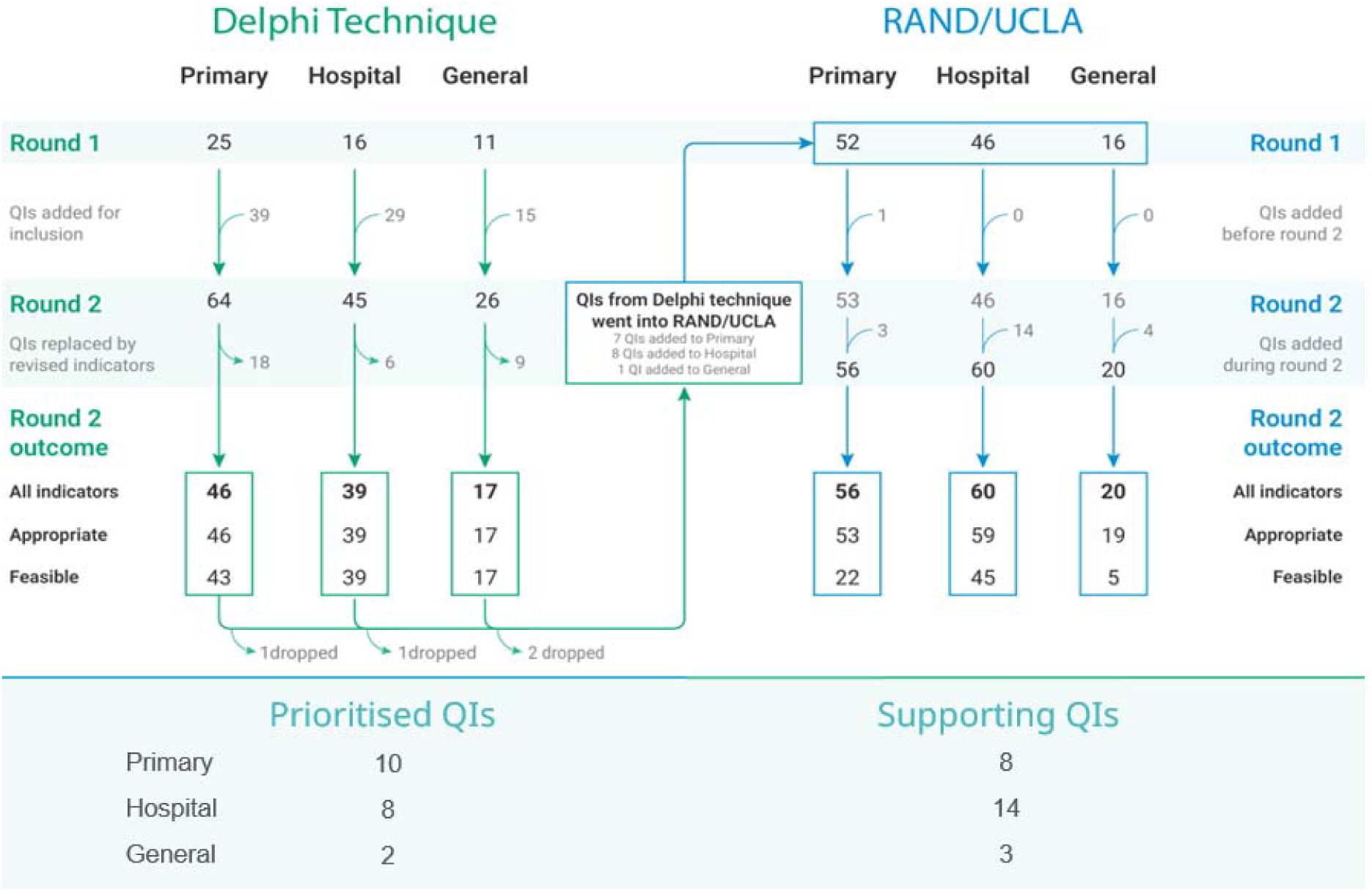
Delphi and RAND/UCLA flowchart showing the number of quality indicators rated per round, the resulting ratings, and the prioritised QI list

Based on the results and suggestions from the panellists in Delphi Round 2, 16 indicators were added to the RAND/UCLA Method (Figure 3). In Round 1 of the RAND/UCLA method, 114 indicators were rated. One new primary care QI was suggested for inclusion in Round 2 resulting in 115 QIs to be rated in RAND/UCLA Round 2 (Figure 3). During the discussion section of Round 2, the panellists added and rated 21 indicators. The final list of indicators after RAND/UCLA Round 2 included 136 QIs.

The QIs resulting from the Delphi and RAND/UCLA methods included primary care, hospital, and general indicators. The primary care and hospital indicators focused on common infections managed in each setting and assessed the appropriateness of antibiotic prescribing in line with AWaRe book guidance, including antibiotic drug choice, dose, and duration. General indicators complemented these infection-specific measures by assessing broader antibiotic use patterns and antimicrobial stewardship practices, providing metrics that can be applied at patient, facility, sub-national, regional, or national levels (Tables S1-S8, pg. 10-54).

### Primary Care

The primary care indicators rated in round 2 of the Delphi Technique resulted in 46 indicators, which were all (100%) rated appropriate (Table S1) while 43 (93.5%) were rated feasible (Table S4) and 3 (6.5%) rated having uncertain feasibility (feasibility rating between 4-6) in a local context. The final list of indicators after Round 2 of the RAND/UCLA method resulted in ratings for 56 Primary Care indicators. Of these 56 indicators, 53 (94.6%) were rated appropriate (Table S1), 22 (39.3%) were rated feasible, and 34 (60.7%) were rated having uncertain feasibility (Table S4) in a global context.

### Hospital Facility

The hospital indicators rate in round 2 of the Delphi Technique resulted in 39 indicators, which were all rated appropriate (Table S2) and feasible (Table S5) on a local level.

Round 2 of the RAND/UCLA method resulted in the rating of 60 hospital indicators, with 59 (98.3%) rated appropriate (Table S2), 45 (75%) rated feasible and 15 (25%) rated having uncertain feasibility in a global context (Table S5).

### General Quality Indicators

There were 17 general indicators rated in round 2 of the Delphi Technique. All the indicators were rated appropriate (Table S3) and feasible (Table S6) in a local context. The RAND/UCLA panellists rated 20 general QIs. Nineteen (95%) of the general QIs were rated appropriate, 5 (25%) were rated feasible and 15 (75%) were rated having uncertain feasibility on a global level (Table S6).

### Prioritised Quality Indicator List

Given the large number of QIs and high appropriateness and feasibility ratings that resulted from the Delphi Technique and RAND/UCLA, a priority list of indicators were developed reflecting measurement of choice of antibiotic along with additional supporting indicators measuring dose and duration of antibiotic treatment (Table 1 & 2). The Primary care priority QIs consisted of 10 indicators and 8 supporting indicator indicators covering 5 infections. The hospital priority QIs included 8 priority indicators and 14 supporting indicators covering 6 infections. The general priority QIs included 2 population level priority indicators and 3 supporting indicators (Table 1 & 2). The prioritisation details are presented in the supplement pg. 55.

**Table 1:**
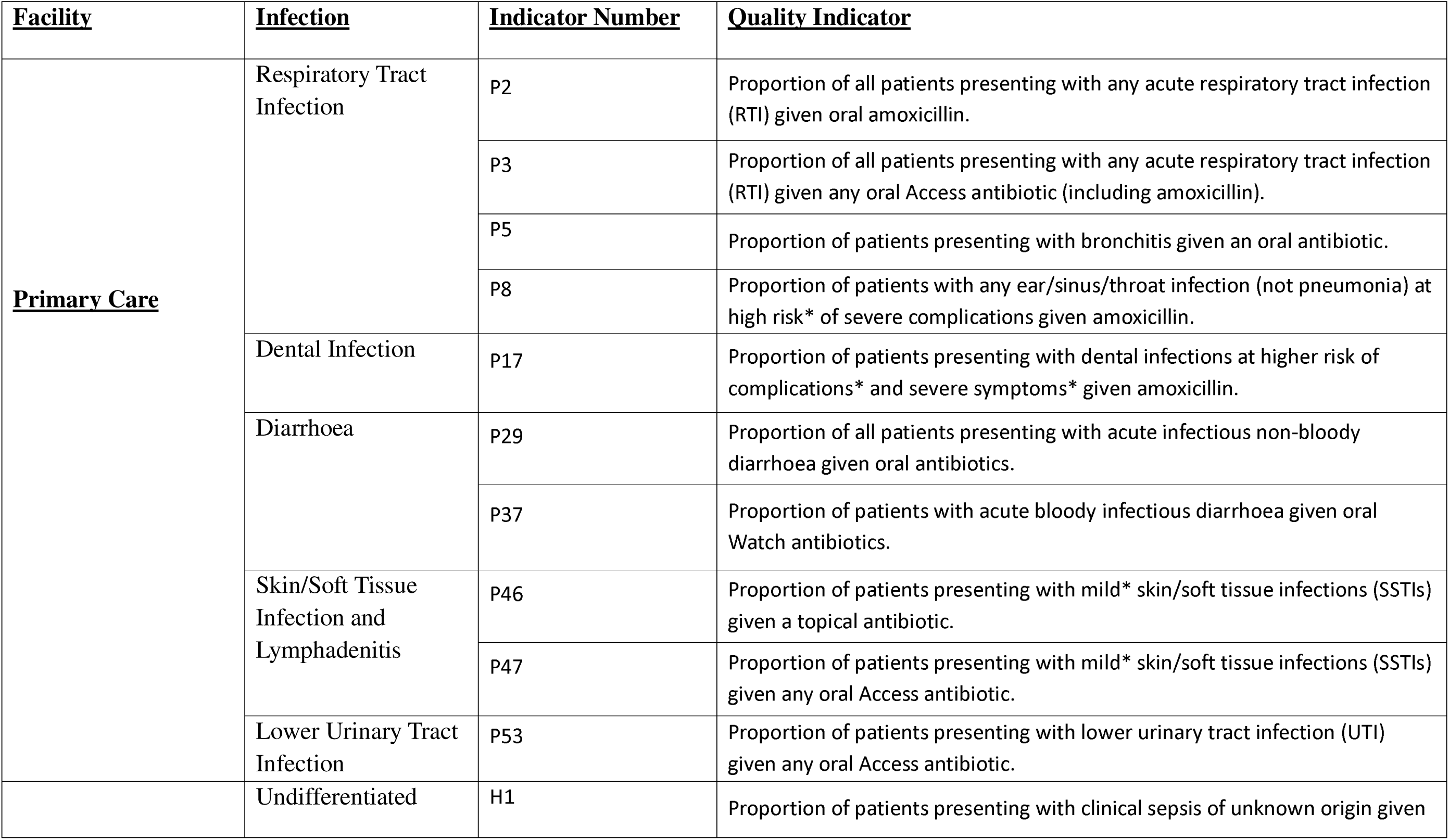
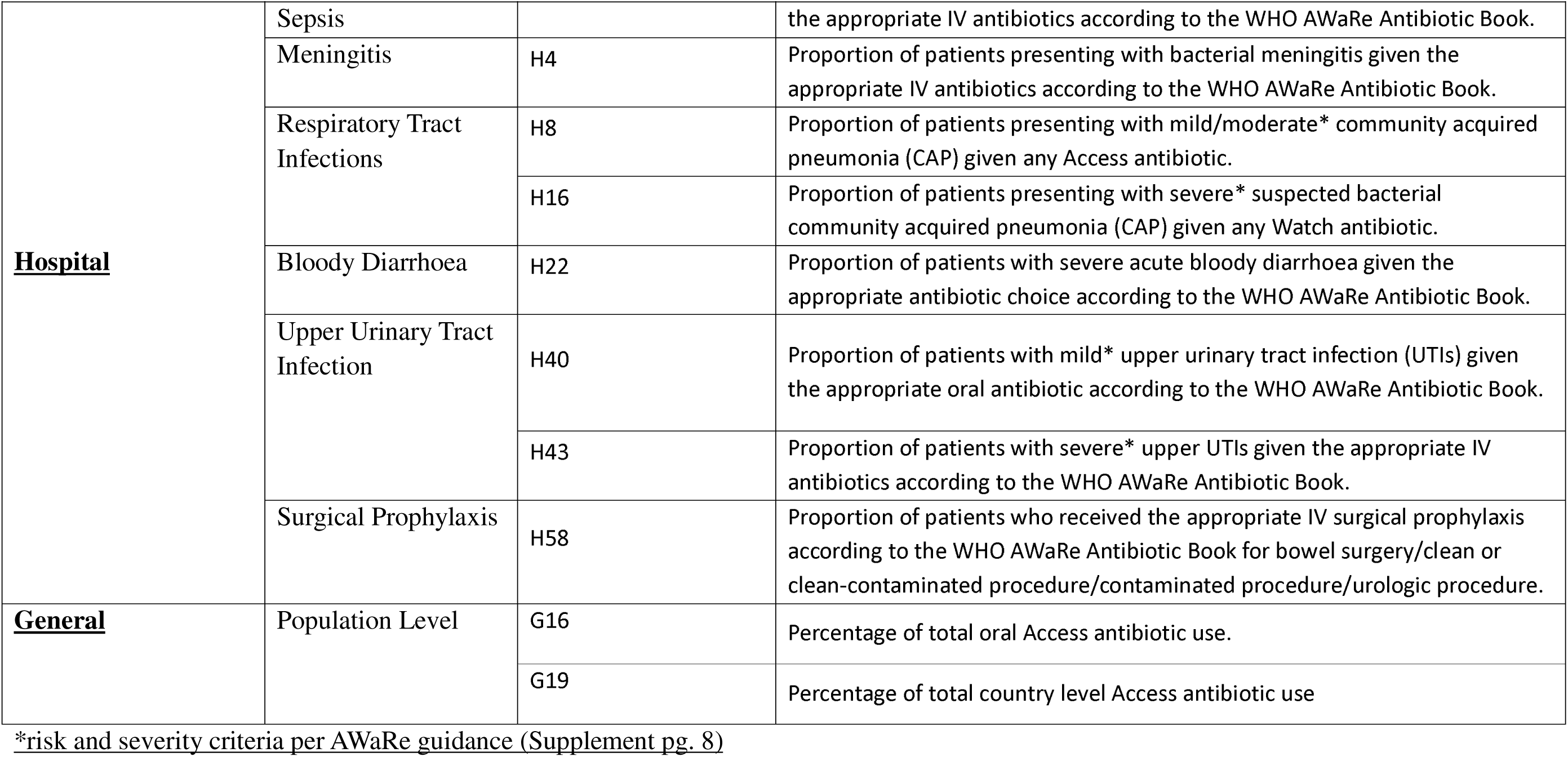
Prioritised List of Quality Indicators: Indicators chosen after prioritisation process reflecting only WHO recommendations and positive indicators.

**Table 2:**
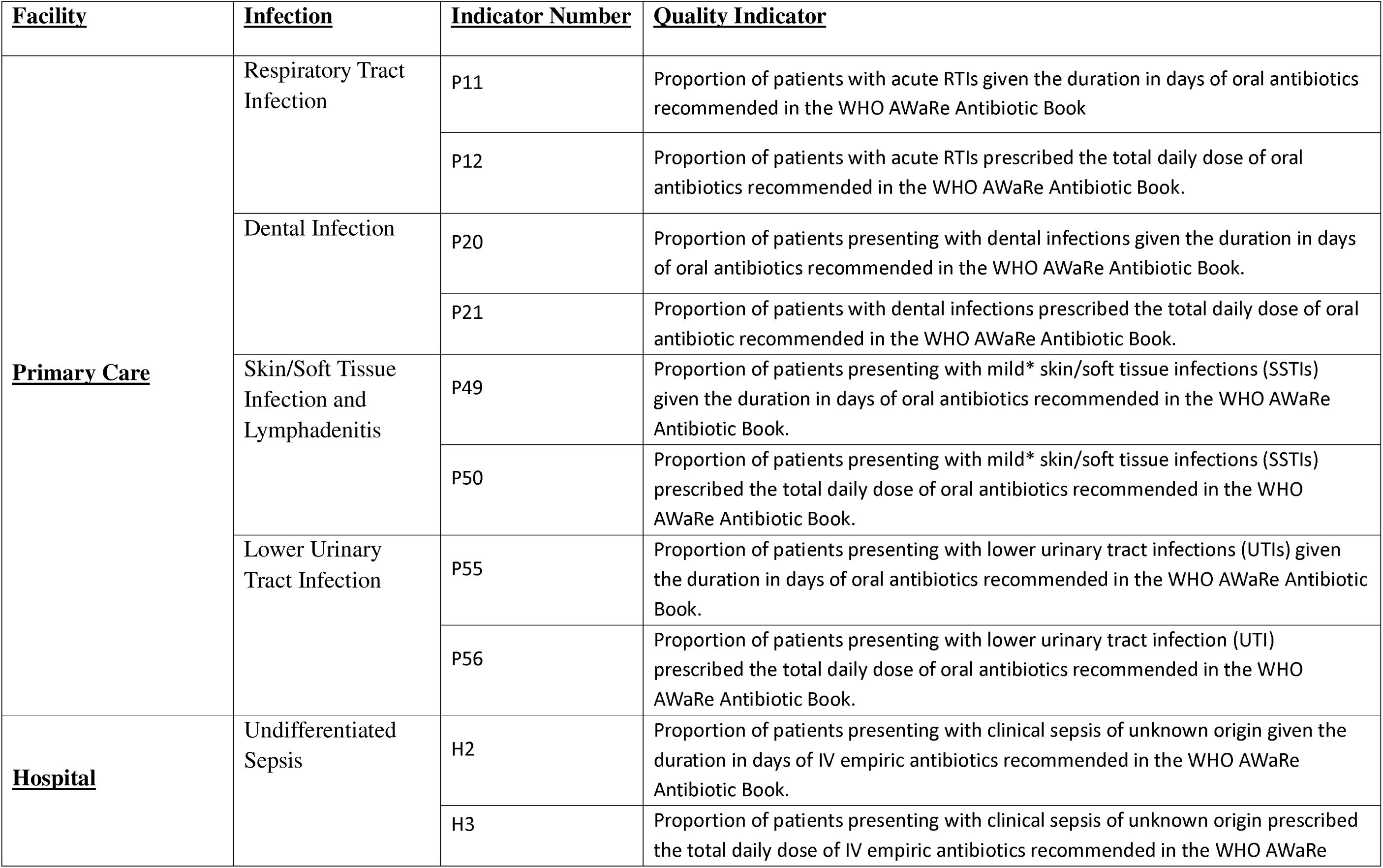
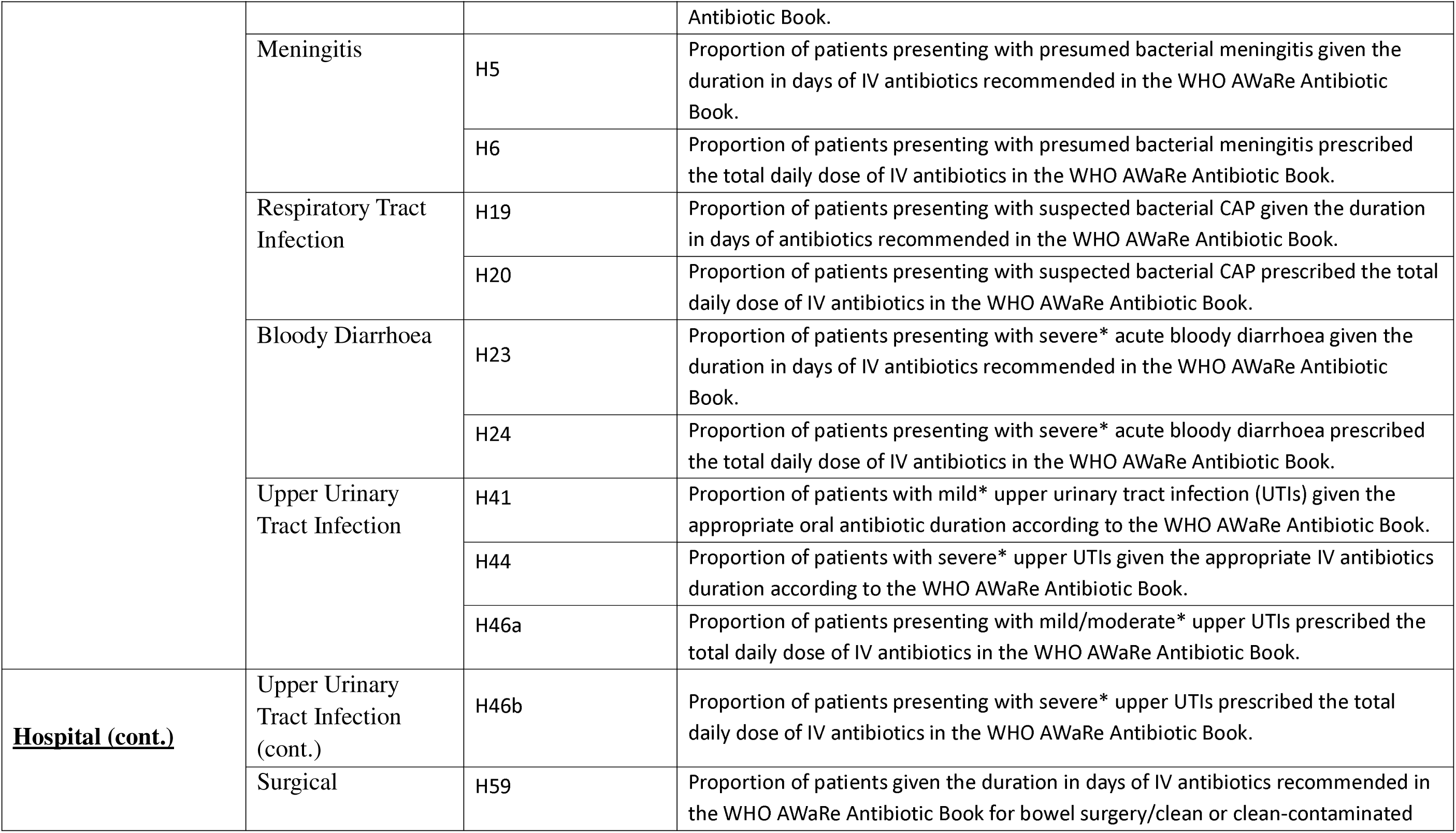
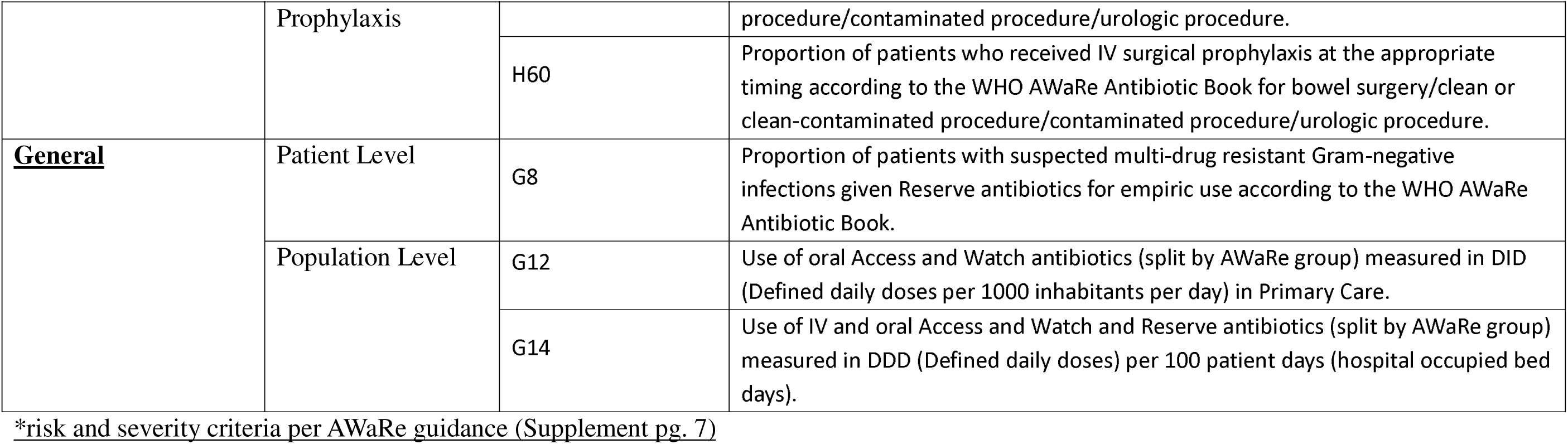
Supporting Quality Indicator list: Indicators chosen through prioritisation process that reflect WHO AWaRe book recommendations of dose and duration of antibiotics.

## Discussion

This study used established consensus methods to develop model sets of quality indicators reflecting WHO AWaRe book recommendations that can be adapted in national and facility level AMS activities to support optimal antibiotic use. The indicators were designed to be practical, feasible, clinically meaningful measures of prescribing quality, reflecting core stewardship principles such as appropriate antibiotic choice, dose, route, and duration, while remaining flexible enough to be implemented using routinely collected data across diverse health system contexts. The iterative process of using both the Delphi Technique and the RAND/UCLA method enabled broad expert input while systematically assessing both clinical evidence, relevance and practical implementation in local and global contexts.

The indicators were highly rated in the Delphi Technique on appropriateness and feasibility in the context of national and local health systems with 102 (100%) of the indicators rated appropriate and 99 (97.1%) currently feasible, in a local context. The RAND/UCLA panellists rated appropriateness and feasibility in a global context and rated 131 indicators (96.3%) appropriate and 72 (52.9%) currently feasible, in a global context.

Given the large number of indicators highly rated during the Delphi Technique and RAND/UCLA method, a subsequent prioritisation exercise was undertaken to develop a more concise set of indicators for feasible local implementation. Indicators selected during the prioritisation process reflect WHO AWaRe book formal recommendations.

Consequently, some indicators that were removed from the prioritised list may remain highly relevant in specific countries, healthcare settings, or patient populations. The prioritised indicators should therefore be considered a model set to support initial implementation and evaluation, with local adaptation based on epidemiology, stewardship priorities, and available human resources and data systems.

One challenge that can hinder the application of quality indicators is the feasibility of collecting accurate data on the numerator and denominator in sufficient numbers of cases, to enable a reliable measurable outcome evaluation over time. ^9,21^ Where QIs are used to compare facilities, appropriate adjustments for case mix and consistent data quality and availability are essential to assess genuine differences in quality of care.

Feedback from the Delphi Technique and the RAND/UCLA panellists highlighted several factors that currently limit the feasibility of some indicators, despite their perceived appropriateness. A substantial proportion of indicators received equivocal feasibility ratings (median 4-6) across both consensus methods, reflecting uncertainty about their routine implementation rather than disagreement about their relevance to improving antibiotic use. These feasibility concerns were largely due to contextual constraints, including variability in data availability, inconsistent clinical documentation or diagnostic capacity, and human and analytical resources required to routinely measure the high number of indicators, particularly in primary care. ^22^

As the indicators were aligned to the AWaRe antibiotic book, they focus predominantly on empiric prescribing, which accounts for the great majority of antibiotic use in the primary care settings. ^14,15^ A recent study that compared 80 national treatment guidelines of paediatric infections in primary care with the AWaRe book, found that most first line recommendations were already closely aligned to AWaRe book recommendations. ^23^ In another primary care study assessing the alignment of adult Standard Treatment Guidelines in 24 countries to the AWaRe book found that many first-line antibiotic treatment recommendations showed moderate alignment with the AWaRe book. ^24^

Our study used QIs identified in a recent narrative review, ^16^ refined them as antibiotic treatment indicators aligned with the AWaRe antibiotic book and developed model indicators for use in primary care and hospital care settings in both high-income countries (HIC) and LMICs, although the emphasis is on improving stewardship interventions in LMICs, especially the primary care settings. ^22,25^ Previous studies have developed a broad range of QIs using similar methodologies, although many of these are relevant to HIC healthcare systems. ^26^ The Driving Reinvestment in Research and Development and Responsible Antibiotic Use (DRIVE-AB) project aimed to identify QIs through a similar method; ^26,27^ however, their indicators were more tailored to high-income countries and placed greater emphasis on microbiology-driven prescribing. ^26^ This can make some of their indicators less feasible in many LMIC settings, where diagnostic capacity may be limited.

The European Surveillance of Antimicrobial Consumption (ESAC) also developed QIs with applicability for Europe. ^28^ ESAC indicators focused on antibiotic prescribing based on national data on antimicrobial use, ^28^ while these AWaRe QIs focus on disease specific patient level data with additional general indicators.

The Delphi Technique and RAND/UCLA panels were selected for regional and gender balance and included diverse health professionals. However, some specialties may not have been represented, and panellists may have had broad subject knowledge but not direct experience with data collection at facility level.

The strengths of this study include a high level of consensus within panels that reflected WHO regional and gender balance, along with high feasibility of collecting data currently, which were included as important attributes for the indicators ^11,19^ in parallel with appropriateness. Both methods closely followed established standards for conducting and reporting consensus studies, ensuring anonymity of ratings, iteration, controlled feedback, statistical stability, with no pressure to force agreement. ^17^

Indicators developed using consensus techniques are evidence based but provide only face-validity rather than empirical validation. ^11,29^ The next step is to evaluate the practical application of these QIs in clinical settings across different countries, including how they can be implemented using routine data, their measurability in practice, and how useful they are for informing decision-making by AMS programs. ^30,31^ Recognising the constraints on data analysis in resource limited settings, a pilot study conducted under a Fleming Fund grant has now developed an automated dashboard tool that uses Global PPS data to measure the AWaRe QIs. ^31^

Addressing the feasibility challenges identified through this work, including strengthening data availability and quality, improving diagnostic capability, and building stewardship capacity, will be essential to enable broader and more consistent use of these indicators. In particular, even where data are available there are limitations related to data quality, incomplete or inconsistent reporting, and lack of standardisation, ^32^ which limit the reliable application and comparability of the QIs. Strengthening routine data systems and expanding the use of electronic and digital data infrastructure, especially among LMICs, will be critical to support accurate measurement and sustainable implementation.

Widespread use of these indicators could lead to improved (1) prescribing quality and outcomes for patients, (2) enhanced monitoring of antibiotic use stratified by AWaRe group and (3) identification of priorities for interventions and policies both globally and locally. For example, a focus on primary care minor respiratory tract infections could be prioritised, as seen in a South African sub-study using the AWaRe Qis, ^33^ which is commonly associated with antibiotic overuse in HIC and LMICs. ^25,32,34^ Optimising the use of antibiotics is a key global priority for better patient care and to reduce the burden of AMR. Achieving this means promoting better locally applied care practices and using measures including QIs to help implement the AWaRe system, ^35^ with its use growing across LMICs to monitor and improve future antibiotic use. ^36^ Standardisation of AWaRe QIs allows sharing of findings and experiences from testing between and within countries to foster a culture of individual and collective transferable learning and quality improvement within and between countries. ^37,38^

## Data Availability

All data produced in the present work are contained in the manuscript and supplementary

## Acknowledgements

Thank you to Baboucarr Njie for providing data management support.

## Members of AWaRe QIs Delphi Panel

Cybele Lara Abad, Aliah Binti Abdul Rahman, Joseph Elikem Efui Acolatse, Faisal Iqbal Afridi, Rahnuma Ahmad, Jameela Al-Salman, Daniel Ankrah, Tehreem Ansari, Siriluck Anunnatsiri, Elizabeth A Ashley, Elizabeth Dodds Ashley, Sadia Asim, Rosmonaliza Asli, Minyon Avent, Lovenish Bains, Laura Barcelona, Sazali Bin Basri, Mirjana Stanić Benić, Adrian John Brink, Franky Buyle, Alessandro Cassini, Lucila Isabel Castro-Pastrana, Michele Cecchini, Sujith J Chandy, Ronald Chitatanga, Hock Hin Chua, Renier Coetzee, Silvia F Costa, Alicia Demirjian, Dhanya Dharmapalan, Daniele Dona, Francis Drobniewski, Lengsea Eng, Cristina Epalza, Jaran Eriksen, Sabiha Yusuf Essack, Joseph Fadare, Ambreen Fatima, Nicholas A Feasey, Moy Siew Fong, Siew Moy Fong, Karen Mary Forrest, Andrew Lofts Gray, Lalit Gupta, Gabriel Levy Hara, Stephan Jürgen Harbarth, Martin Hojman, Muhammad Akram Hossain, Sigurd Høye, Li Yang Hsu, Vu Thi Lan Huong, Brecht Ingelbeen, Elias Iosifidis, Yasmina Johnson, Thomas Kesteman, Faryal Khamis, Felix Khuluza, Hong Bin Kim, Ioannis Kopsidas, Marlieke E.A. de Kraker, David Kulapani, Nishant Kumar, Deepak Kumar, Direk Limmathurotsakul, Lowell Ling, Po-Yu Liu, Mark Loeb, Afzalunnessa Binte Lutfor, Syed Faisal Mahmood, Amos Yared Massele, Matshediso Matome, Mohd Zulfakar Mazlan, Aalaa Moghith, Jamil Muqtadir, Zia Ul Mustafa, Behzad Nadjm, Nosheen Nasir, Erni Juwita Nelwan, Olaf Neth, Rong Xiang Ng, Thuy Thi Phuong Nguyen, Mi Budiongo Yave Ntondele, Christina W. Obiero, Jaap Ten Oever, Olayinka Olabode Ogunleye, Ioana Diana Olaru, Margaret Ngwono Oluka, Mong How Ooi, Helena Owusu, Ayodhia Pitaloka Pasaribu, Mical Paul, Nuno Rocha Pereira, Rongpong Plongla, Sasheela Ponnampalavanar, Ana Paula Matos Porto, Gabriela Jana Prutsky, Céline Pulcini, Keshav Rai, Glaiza Palaganas Ranges, Amrita Ronnachit, Nirmeen Ahmed Sabry, Zeti Norfidiyati Salmuna, Monira Sarmin, Luigia Scudeller, Nour Shamas, Hira Zafar Siddiqui, Rafael Almeida da Silva, Robert Sinto, Jyoti Somani, Helmi Sulaiman, Cindy Shuan Ju Teh, Xun Ting Tiong, Chitra Tomy, Deborah Tong, Paul Torpiano, Chien-Hao Tseng, Paul Turner, Yuki Uehara, Ann Versporten, Vera Vlahović-Palčevski, Connie Walyaro, Heiman Frank Louis Wertheim, Phoebe CM Williams, Sumudu Suranandee Yattowita Withanage, Yonghong Xiao, S. M. Sabrina Yesmin, Ghada Ali Mohamed Younis, Veronica Zanichelli

## Author Contributions

MS, SMC, AC conceptualised the study

EF, GL conducted narrative review to establish the initial list of quality indicators

AC, JG, BG, CEM, SMC, MS adapted indicators before and after panel ratings

CLA, JEEA, FIA, RA, JA, DA, TA, SA, EAA, EDA, SA, RA, MA, LB, LB, SBB, MSB, AJB, FB, AC, LIC, MC, SJC, RC, HHC, RC, SFC, AD, DD, DD, FD, LE, CE, JE, SYE, JF, AF, NAF, MSF, SMF, KMF, ALG, LG, GLH, SJH, MH, MAH, SH, LYH, VTLH, BI, EI, YJ, TK, FK, FK, HBK, IK, MEDK, DK, NK, DK, DL, LL, PL, ML, ABL, SFM, AYM, MM, MZM, AM, JM, ZUM, BN, NN, EJN, ON, RXN, TTPN, MBYN, CWO, JTO, OOO, IDO, MNO, MHO, HO, APP, MP, NRP, RP, SP, APMP, GJP, CP, ABAR, KR, GPR, AR, NAS, ZNS, MS, LS, NS, HZS, RADS, RS, JS, HS, CSJT, XTT, CT, DT, PT, CT, PT, YU, AV, VV, CW, HFLW, PCW, SSYW, YX, SMSY, GAMY, VZ rated indicators as part of the Delphi Panel

PC, CO, NTTD, ET, EW, JAS, BH, YMK, ZS, SG, FCL, MM rated indicators as part of the RAND/UCLA Panel

JG managed the panellists and data

AH, FD, JG analysed the indicators and consensus ratings

AH, JG, SMC wrote the first draft of the manuscript

All authors critically reviewed and revised the manuscript

## Transparency Declaration

The authors have no conflict of interest to declare. The findings and conclusions in this report are those of the authors and do not necessarily represent the official position of WHO or any of the institutions mentioned.

Ethical approval for this study was obtained from St. George’s, University of London Research Ethics Committee under the ADILA project (REC reference: 2022.0113)

