## Supplemental File for "Development of AWaRe quality indicators for optimal antibiotic use"

‡AWaRe QIs Delphi Panel *(see full list of authors in acknowledgment section)*

§ RAND/UCLA Panel

Authors apart from first and last are listed in alphabetical order

*Table S5: Feasibility Ratings of Hospital model Quality Indicators Rated in the Delphi Technique and RAND/UCLA Panel ........* ***Error! Bookmark not defined.***

### 1. Delphi Technique and RAND/UCLA method Detailed Process

In both consensus techniques, the panellists were sent an online MS Excel® spreadsheet and instructions by email under three classification headings: primary care, hospital facility, and general indicators

To facilitate ratings in Round 2 of each method, panel members received a summary of their individual ratings from Round 1 and the median panel rating for each indicator, as well as the level of consensus (Agreement, Disagreement, Equivocal) (Figure S1).

*Data Analysis*

The median rating for each indicator was calculated for clarity and appropriateness respectively in Round 1 and feasibility and appropriateness in Round 2. [1] Supplement Figure 1 shows the process and definition of the different levels of agreement by which the indicators were analysed. [2,3]

#### Box S1: Number of Panellists in Delphi Technique and RAND/UCLA Method distributed by WHO Region

| WHO Region[1] | Delphi Round 1 n (%) | Delphi Round 2 n (%) | RAND/UCLA Round 1 & 2 n (%) |
|---|---|---|---|
| AFR | 17 (16.3%) | 17 (15.9%) | 1 (8.3%) |
| AMR | 7 (6.7%) | 8 (7.5%) | 3 (25.0%) |
| EMR | 7 (6.7%) | 13 (12.1%) | 2 (16.7%) |
| EUR | 25 (24%) | 24 (22.4%) | 3 (25.0%) |
| SEAR | 21 (20.2%) | 18 (16.8%) | 2 (16.7%) |
| WPR | 27 (26.0%) | 27 (25.2%) | 1 (8.3%) |
| **TOTAL** | 104 | 107 | 12 |

[1] AFR: African Region, AMR: Region of the Americas, EMR: Eastern Mediterranean Region, EUR: European Region, SEAR: South-East Asia Region, WPR: Western Pacific Region

#### Box S2: 9-point integer scales used in Delphi Technique and RAND/UCLA Appropriateness Method

| Rating | Clarity: round 1 | Appropriateness: rounds 1 & 2 | Feasibility: round 2 |
|---|---|---|---|
| **1** | Very unclear and ambiguous | Completely inappropriate everywhere: NO exceptions | Completely infeasible everywhere: NO exceptions |
| **2** | Unclear and ambiguous | Generally inappropriate – rare exceptions | Generally infeasible – rare exceptions |
| **3** | Moderately unclear and ambiguous | Generally inappropriate - some regional or continent exceptions | Generally infeasible- some zonal or continent exceptions |
| **4** | Slightly unclear and ambiguous | Equivocal – generally inappropriate even though appropriate in many areas/ continents | Equivocal – generally infeasible even though feasible in many areas/ continents |
| **5** | Neither clear nor unclear | Equivocal – 50:50 | Equivocal – 50:50 |
| **6** | Slightly clear and unambiguous | Equivocal – generally appropriate even though inappropriate in some areas/ continents | Equivocal – generally feasible even though infeasible in some areas/continents |
| **7** | Moderately clear and unambiguous | Generally appropriate - some regional or continent exceptions | Generally feasible - some zonal or continent exceptions |
| **8** | Clear and unambiguous | Generally appropriate – rare exceptions | Generally feasible – rare exceptions |
| **9** | Very clear and unambiguous | Completely appropriate everywhere: NO exceptions | Completely feasible everywhere: NO exceptions |

Figure S1: Definitions of clarity, appropriateness and feasibility measurements used in Delphi Technique and RAND/UCLA Appropriateness Method

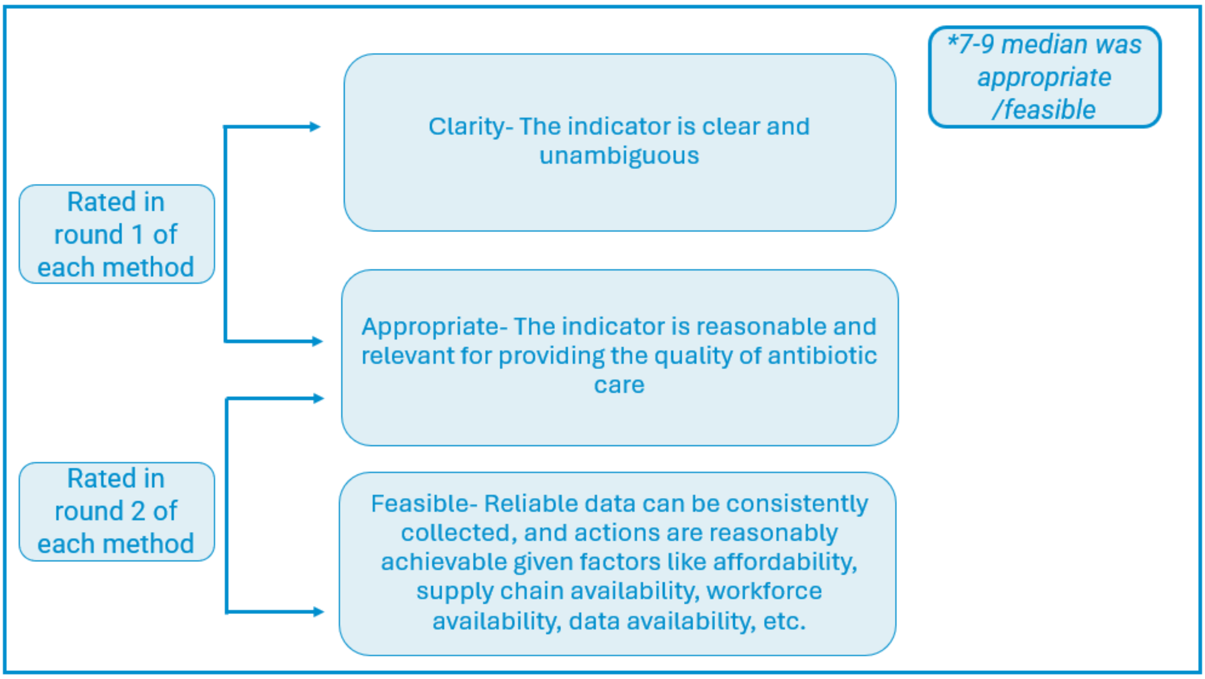

Figure S2: Levels of agreement/consensus in the Delphi Technique and RAND/UCLA processes

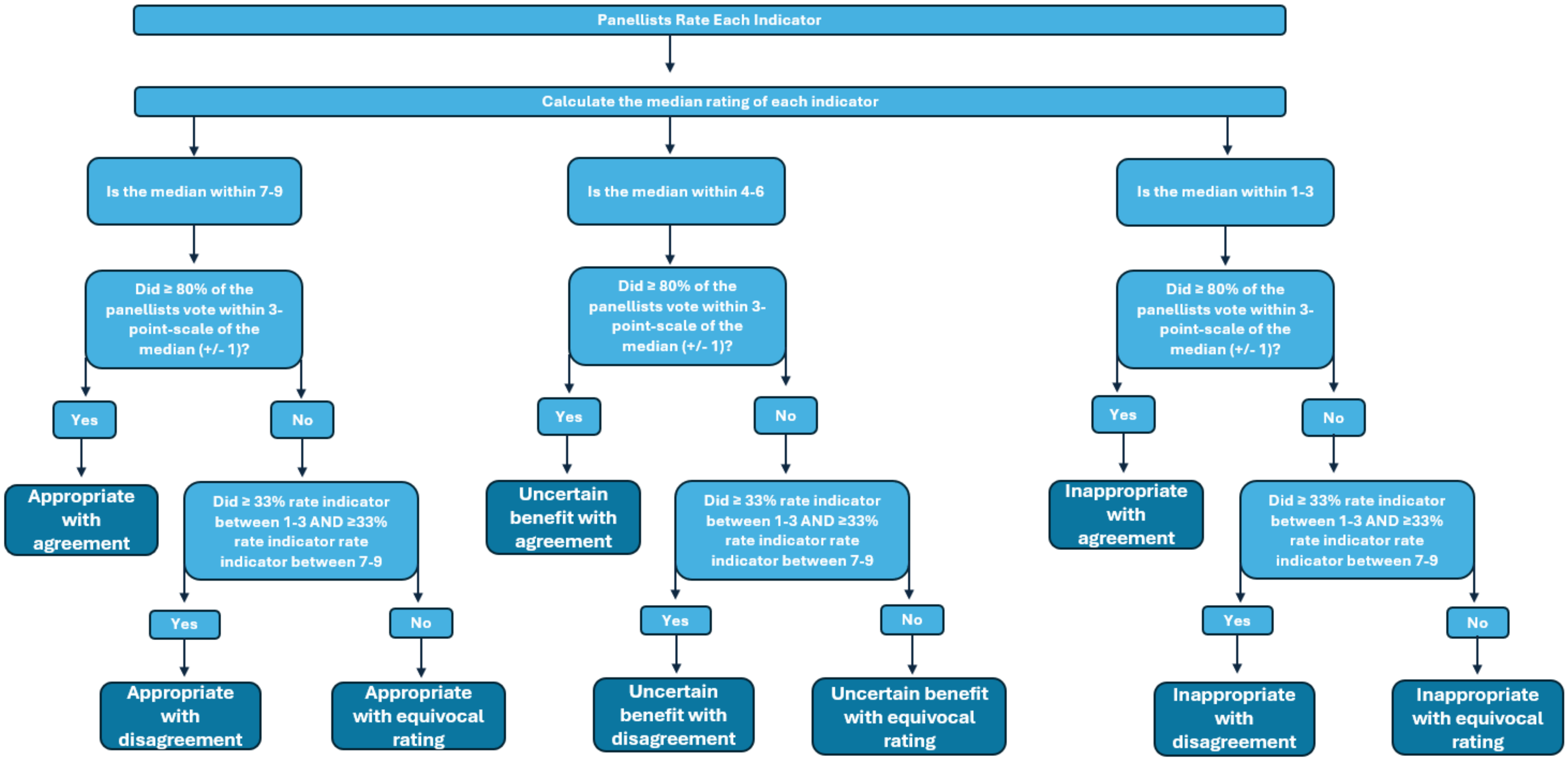

#### 2. Severity and Risk Criteria According to WHO AWaRe book [4]

The quality indicators include several common conditions, and some indicators refer to mild/moderate or severe conditions as well as high risk patients. The following criteria can be used to assess what mild/moderate to severe infections look like and who are high risk patients.

##### Primary Care infection risk factors

1. Community Acquired Pneumonia (AWaRe book pg. 147)
    a. Undernutrition, HIV infection common risk factors for severe infection in children under 5
    b. Other severe comorbid illnesses
2. Diarrhoea (pg. 169, 185)
    a. Co-existing conditions (e.g. pregnancy)
    b. Comorbidities/ immunocompromised patients
    c. Severe malnutrition

##### Primary Care Mild Infection Symptoms

1. Skin/Soft Tissue infection (AWaRe book pg. 193)
    a. Superficial bacterial skin infection, not affecting deeper tissue layers

##### Hospital Severe Infection Symptoms

1. Community Acquired Pneumonia (AWaRe book pg. 362)
    a. Higher short-term mortality risk and need for hospital/intensive care admission
    b. CURB-65 greater or equal to 2
    c. In children: fast breathing with or without chest indrawing, other danger signs such as inability to breastfeed or drink, convulsions, lethargy, or reduced level of consciousness
    d. Inability to maintain oral therapy (adult)
2. Upper Urinary Tract Infection (AWaRe book pg. 474)
    a. Nausea/vomiting
    b. High grade fever

c. Severe flank pain
d. Severe costovertebral angle tenderness
e. Complicated UTI
f. Patients who are critically ill, with sepsis and/or septic shock

3. Diarrhoea (AWaRe book pg. 169, 185)
    a. Critically ill with confirmed/ suspected intestinal perforation, peritonitis, sepsis, or septic shock
    b. Severe dehydration
    c. Lethargic and/or unconscious
    d. Sunken eyes
    e. Inability to drink
    f. Skin pinch goes back very slowly (2 or more seconds)
4. Intra-abdominal infection (AWaRe book pg. 396, 414, 434, 454)
    a. Critically ill with signs of sepsis or septic shock
    b. Hypotension and signs of organ hypoperfusion may be present in cases of organ failure

#### 3. Delphi Technique and RAND/UCLA Appropriateness and Feasibility Results

##### Table S1: Appropriateness Ratings of Primary Care model QIs Rated in the Delphi Technique and RAND/UCLA Panel

| Primary Care Indication | Indicator list | Quality Indicator | Delphi Approp Median[1] | Delphi Approp Consensus[2] | Delphi Percent Agreement (n=107)[3] | RAND/UCLA Approp Median[1] | RAND/UCLA Approp Consensus[2] | RAND/UCLA Percent Agreement (n=12)[3] |
|---|---|---|---|---|---|---|---|---|
| **Respiratory Tract Infections** | P1 | Proportion of all patients presenting with any acute respiratory tract infection (RTI) given an oral antibiotic. | 8 | E | 78.5% | 8 | A | 83.3% |
| | P2 | Proportion of all patients presenting with any acute respiratory tract infection (RTI) given oral amoxicillin. | 7 | E | 60.8% | 8 | A | 91.7% |
| | P3 | Proportion of all patients presenting with any acute respiratory tract infection (RTI) given any oral Access antibiotic (including amoxicillin). | 8 | E | 76.6% | 8 | A | 91.7% |
| | P4 | Proportion of all patients presenting with any acute respiratory tract infection (RTI) given any oral Watch antibiotic. | 8 | E | 76.6% | 8 | A | 91.7% |
| | P5 | Proportion of patients presenting with bronchitis given an oral antibiotic. | 7 | E | 70.8% | 7 | E | 75% |
| | P6 | Proportion of patients with any ear/sinus/throat infection (not pneumonia) given an oral antibiotic. | 8 | E | 78.5% | 8 | A | 83.3% |
| | P7 | Proportion of patients with any ear/sinus/throat infection (not pneumonia) at high risk* of severe complications given an oral antibiotic. | 8 | E | 78.5% | 8 | A | 100% |
| | P8 | Proportion of patients with any ear/sinus/throat infection (not pneumonia) at high risk* of severe complications given amoxicillin. | 7 | E | 68.9% | 7 | A | 91.7% |

| Primary Care Indication | Indicator list | Quality Indicator | Delphi Approp Median[1] | Delphi Approp Consensus[2] | Delphi Percent Agreement (n=107)[3] | RAND/UCLA Approp Median[1] | RAND/UCLA Approp Consensus[2] | RAND/UCLA Percent Agreement (n=12)[3] |
|---|---|---|---|---|---|---|---|---|
| **Respiratory Tract Infections (cont.)** | P9 | Proportion of patients with any ear/sinus/throat infection (not pneumonia) at high risk* of severe complications given any oral Access antibiotic (including amoxicillin). | 8 | A | 82.2% | 7 | A | 91.7% |
| | P10 | Proportion of patients at lower risk of a bacterial respiratory tract infection given an oral antibiotic. | 7 | E | 69.2% | NA | NA | NA |
| | P11 | Proportion of patients with acute RTIs given the duration in days of oral antibiotics recommended in the WHO AWaRe antibiotic book | 8 | E | 73.8% | 7 | A | 83.3% |
| | P12 | Proportion of patients with acute RTIs prescribed the total daily dose of oral antibiotics recommended in the WHO AWaRe antibiotic book. | NA | NA | NA | 7 | E | 75% |
| | P13 | Proportion of patients with bacterial RTIs given oral Access or Watch antibiotics. | 8 | E | 79.4% | 7 | E | 75% |
| **Dental Infections** | P14 | Proportion of otherwise healthy adults presenting with dental infections given an oral antibiotic. | 7 | E | 68.2% | 8 | E | 75% |
| | P15 | Proportion of otherwise healthy adults presenting with severe* dental infections given an oral antibiotic. | 7 | E | 72.9% | 8 | A | 83.3% |
| | P16 | Proportion of patients presenting with dental infections at high risk* of severe complications given an oral antibiotic. | 7 | E | 73.8% | 8 | E | 75% |
| | P17 | Proportion of patients presenting with dental infections at high risk* of severe complications given amoxicillin. | 7 | E | 70.1% | 7 | A | 83.3% |
| | P18 | Proportion of patients presenting with dental infections at high risk* of severe complications given any oral Access antibiotic (including amoxicillin). | 7 | E | 74.8% | 8 | E | 75% |

| Primary Care Indication | Indicator list | Quality Indicator | Delphi Approp Median[1] | Delphi Approp Consensus[2] | Delphi Percent Agreement (n=107)[3] | RAND/UCLA Approp Median[1] | RAND/UCLA Approp Consensus[2] | RAND/UCLA Percent Agreement (n=12)[3] |
|---|---|---|---|---|---|---|---|---|
| **Dental Infections (cont.)** | P19 | Proportion of patients with lower risk (of bacterial infection) dental infections given oral antibiotics. | 7 | E | 68.2% | 7 | A | 91.7% |
| | P20 | Proportion of patients presenting with dental infections given the duration in days of oral antibiotics recommended in the WHO AWaRe antibiotic book. | 8 | A | 82.1% | 7 | A | 83.3% |
| | P21 | Proportion of patients with dental infections prescribed the total daily dose of oral antibiotic recommended in the WHO AWaRe antibiotic book. | NA | NA | NA | 7 | E | 75% |
| | P22 | Proportion of patients with dental infections given oral Watch antibiotics. | 8 | A | 84.1% | 7 | A | 83.3% |
| **Bacterial Eye Infection** | P23 | Proportion of patients presenting with bacterial eye infections given the duration in days of oral antibiotics recommended in the WHO AWaRe antibiotic book. | 7 | E | 74.8% | 7 | E | 58.3% |
| | P24 | Proportion of patients with bacterial eye infections prescribed the total daily dose of oral antibiotics recommended in the WHO AWaRe antibiotic book. | NA | NA | NA | 6 | E | 58.3% |
| | P25 | Proportion of patients with eye infections prescribed the total daily dose of oral antibiotics recommended in the WHO AWaRe antibiotic book | NA | NA | NA | 5 | D | 33.3% |
| | P26 | Proportion of patients presenting with eye infection given an oral antibiotic. | 8 | E | 71.0% | 8 | A | 83.3% |
| | P27 | Proportion of patients presenting with eye infection at high risk* of severe complications given any oral Access antibiotic. | 8 | E | 74.8% | 7 | A | 100% |
| | P28 | Proportion of patients presenting with eye infection at high risk* of severe complications given any oral Watch antibiotic. | 7 | E | 67.3% | 7 | A | 91.7% |

| Primary Care Indication | Indicator list | Quality Indicator | Delphi Approp Median[1] | Delphi Approp Consensus[2] | Delphi Percent Agreement (n=107)[3] | RAND/UCLA Approp Median[1] | RAND/UCLA Approp Consensus[2] | RAND/UCLA Percent Agreement (n=12)[3] |
|---|---|---|---|---|---|---|---|---|
| **Diarrhoea** | P29 | Proportion of all patients presenting with acute infectious non-bloody diarrhoea given oral antibiotics. | 8 | A | 81.3% | 8 | E | 66.7% |
| | P30 | Proportion of all patients presenting with high-risk* acute infectious non-bloody diarrhoea given any oral Access antibiotic. | 8 | E | 79.4% | 8 | A | 83.3% |
| | P31 | Proportion of all patients presenting with high-risk* acute infectious non-bloody diarrhoea given any oral Watch antibiotic. | 8 | E | 77.6% | 7 | A | 83.3% |
| | P32 | Proportion of otherwise healthy patients presenting with acute infectious non-bloody diarrhoea given an oral antibiotic. | 8 | E | 72.0% | 7 | E | 75% |
| | P33 | Proportion of patients presenting with high-risk* acute infectious non-bloody diarrhoea given the duration in days of oral antibiotics recommended in the WHO AWaRe antibiotic book. | 8 | E | 73.8% | 7 | A | 83.3% |
| | P34 | Proportion of patients presenting with high-risk* acute infectious non-bloody diarrhoea prescribed the total daily dose of oral antibiotics recommended in the WHO AWaRe antibiotic book. | NA | NA | NA | 7 | E | 75% |
| | P35 | Proportion of patients with acute bloody infectious diarrhoea given oral Access or Watch antibiotics. | 8 | A | 80.4% | 7 | E | 75% |
| | P36 | Proportion of patients with acute bloody infectious diarrhoea given oral Access antibiotics. | NA | NA | NA | 8 | A | 83.3% |
| | P37 | Proportion of patients with acute bloody infectious diarrhoea given oral Watch antibiotics. | NA | NA | NA | 8 | A | 100% |
| **Skin/Soft Tissue Infections and Lymphadenitis** | P38 | Proportion of patients presenting with acute lymphadenitis given an oral antibiotic. | 7 | E | 66.3% | 7 | E | 75% |

| Primary Care Indication | Indicator list | Quality Indicator | Delphi Approp Median[1] | Delphi Approp Consensus[2] | Delphi Percent Agreement (n=107)[3] | RAND/UCLA Approp Median[1] | RAND/UCLA Approp Consensus[2] | RAND/UCLA Percent Agreement (n=12)[3] |
|---|---|---|---|---|---|---|---|---|
| **Skin/Soft Tissue Infections and Lymphadenitis (cont.)** | P39 | Proportion of patients presenting with higher risk* (of bacterial infection) acute lymphadenitis given an oral antibiotic. | 7 | E | 74.5% | 7 | A | 83.3% |
| | P40 | Proportion of patients presenting with higher risk* (of bacterial infection) acute lymphadenitis given any oral Access antibiotic. | 7 | E | 72.9% | 7 | A | 91.7% |
| | P41 | Proportion of patients presenting with higher risk* (of bacterial infection) acute lymphadenitis given any oral Watch antibiotic. | 7 | E | 70.1% | 7 | A | 83.3% |
| | P42 | Proportion of patients with lower risk (of bacterial infection) acute lymphadenitis given oral antibiotics. | 7 | E | 63.6% | 7 | E | 75% |
| | P43 | Proportion of patients presenting with acute lymphadenitis given the duration in days of oral antibiotics recommended in the WHO AWaRe antibiotic book. | 8 | E | 69.1% | 7 | E | 75% |
| | P44 | Proportion of patients presenting with acute lymphadenitis prescribed the total daily dose of oral antibiotics recommended in the WHO AWaRe antibiotic book. | NA | NA | NA | 6 | E | 66.7% |
| | P45 | Proportion of patients presenting with mild* skin/soft tissue infections (SSTIs) given an oral antibiotic. | 8 | A | 80.4% | 7 | A | 100% |
| | P46 | Proportion of patients presenting with mild* skin/soft tissue infections (SSTIs) given a topical antibiotic. | NA | NA | NA | 7 | A | 83.3% |
| | P47 | Proportion of patients presenting with mild* skin/soft tissue infections (SSTIs) given any oral Access antibiotic. | 8 | A | 82.2% | 7 | A | 91.7% |

| Primary Care Indication | Indicator list | Quality Indicator | Delphi Approp Median[1] | Delphi Approp Consensus[2] | Delphi Percent Agreement (n=107)[3] | RAND/UCLA Approp Median[1] | RAND/UCLA Approp Consensus[2] | RAND/UCLA Percent Agreement (n=12)[3] |
|---|---|---|---|---|---|---|---|---|
| **Skin/Soft Tissue Infections and Lymphadenitis (cont.)** | P48 | Proportion of patients presenting with mild* skin/soft tissue infections (SSTIs) given any oral Watch antibiotic. | 8 | A | 81.3% | 7 | A | 91.7% |
| | P49 | Proportion of patients presenting with mild* skin/soft tissue infections (SSTIs) given the duration in days of oral antibiotics recommended in the WHO AWaRe antibiotic book. | 8 | A | 81.1% | 7 | A | 100% |
| | P50 | Proportion of patients presenting with mild* skin/soft tissue infections (SSTIs) prescribed the total daily dose of oral antibiotics recommended in the WHO AWaRe antibiotic book. | NA | NA | NA | 7 | A | 100% |
| **Lower Urinary Tract Infections** | P51 | Proportion of low-risk patients* with positive urine test (positive urine leucocytes/leucocyte esterase or positive urine culture), but no urinary tract infection (UTI) symptoms (e.g., no dysuria, no increased urinary urgency and frequency, no lower abdominal pain or discomfort or sometimes visible haematuria), given oral antibiotics. | 8 | E | 74.8% | 7 | E | 66.7% |
| | P52 | Proportion of patients presenting with lower urinary tract infection (UTI) given an oral antibiotic. | 8 | E | 79.5% | 8 | A | 91.7% |
| | P53 | Proportion of patients presenting with lower urinary tract infection (UTI) given any oral Access antibiotic. | 8 | A | 86.0% | 8 | A | 91.7% |
| | P54 | Proportion of patients presenting with lower urinary tract infection (UTI) given any oral Watch antibiotic. | 8 | A | 86.9% | 8 | A | 83.3% |
| | P55 | Proportion of patients presenting with lower urinary tract infections (UTIs) given the duration in days of oral antibiotics recommended in the WHO AWaRe antibiotic book. | 8 | A | 85.0% | 7 | A | 83.3% |

| Primary Care Indication | Indicator list | Quality Indicator | Delphi Approp Median[1] | Delphi Approp Consensus[2] | Delphi Percent Agreement (n=107)[3] | RAND/UCLA Approp Median[1] | RAND/UCLA Approp Consensus[2] | RAND/UCLA Percent Agreement (n=12)[3] |
|---|---|---|---|---|---|---|---|---|
| **Lower Urinary Tract Infections (cont.)** | P56 | Proportion of patients presenting with lower urinary tract infection (UTI) prescribed the total daily dose of oral antibiotics recommended in the WHO AWaRe antibiotic book. | NA | NA | NA | 7 | A | 100% |
| | P57 | Proportion of patients with lower urinary tract infection (UTIs) given oral Watch antibiotics. | 8 | A | 85.0% | 7 | E | 75% |

[1] Panellist Median Rating on scale of 1-9
[2] Consensus Rating: A=Agreement, E=Equivocal, D=Disagreement, NA= Indicator not rated in round
[3] Percent of panellists that rated within the three-point scale of the median
*Risk criteria as per AWaRe guidance
*Severity criteria as per AWaRe guidance

Table S2: Appropriateness ratings of Hospital Model QIs Rated in the Delphi Technique and RAND/UCLA Panel

| Hospital Indication | Indicator list | Quality Indicator | Delphi Approp Median[1] | Delphi Approp Consensus[2] | Delphi Percent Agreement (n=107)[3] | RAND/UCLA Approp Median[1] | RAND/UCLA Approp Consensus[2] | RAND/UCLA Percent Agreement (n=12)[3] |
|---|---|---|---|---|---|---|---|---|
| **Undifferentiated Sepsis** | H1 | Proportion of patients presenting with clinical sepsis of unknown origin given the appropriate IV antibiotics according to the WHO AWaRe antibiotic book. | 8 | A | 83.2% | 8 | A | 91.7% |
| | H2 | Proportion of patients presenting with clinical sepsis of unknown origin given the duration in days of IV empiric antibiotics recommended in the WHO AWaRe antibiotic book. | 8 | A | 81.3% | 8 | A | 91.7% |
| | H3 | Proportion of patients presenting with clinical sepsis of unknown origin prescribed the total daily dose of IV empiric antibiotics recommended in the WHO AWaRe antibiotic book. | NA | NA | NA | 7 | A | 91.7% |
| **Meningitis** | H4 | Proportion of patients presenting with bacterial meningitis given the appropriate IV antibiotics according to the WHO AWaRe antibiotic book. | 8 | A | 90.7% | 8 | A | 100% |
| | H5 | Proportion of patients presenting with presumed bacterial meningitis given the duration in days of IV antibiotics recommended in the WHO AWaRe antibiotic book. | 8 | A | 90.7% | 8 | A | 91.7% |
| | H6 | Proportion of patients presenting with presumed bacterial meningitis prescribed the total daily dose of IV antibiotics in the WHO AWaRe antibiotic book. | NA | NA | NA | 7 | A | 91.7% |
| **Respiratory Tract Infections** | H7 | Proportion of patients presenting with non-severe* community acquired pneumonia (CAP) given amoxicillin. | 8 | E | 77.6% | 8 | A | 83.3% |

| Hospital Indication | Indicator list | Quality Indicator | Delphi Approp Median[1] | Delphi Approp Consensus[2] | Delphi Percent Agreement (n=107)[3] | RAND/UCLA Approp Median[1] | RAND/UCLA Approp Consensus[2] | RAND/UCLA Percent Agreement (n=12)[3] |
|---|---|---|---|---|---|---|---|---|
| **Respiratory Tract Infections (cont.)** | H8 | Proportion of patients presenting with non-severe* community acquired pneumonia (CAP) given any Access antibiotic. | 8 | A | 83.2% | 8 | E | 75% |
| | H9 | Proportion of patients presenting with non-severe* community acquired pneumonia (CAP) given any Watch antibiotic. | 8 | E | 78.5% | 8 | A | 83.3% |
| | H10 | Proportion of patients with severe bacterial community acquired pneumonia (CAP) given the appropriate IV/oral antibiotic (drug choice and duration) according to the WHO AWaRe antibiotic book. | 8 | A | 87.8% | 8 | A | 91.7% |
| | H11 | Proportion of patients with severe bacterial CAP given the appropriate IV/oral antibiotic drug according to the WHO AWaRe antibiotic book. | NA | NA | NA | 8 | A | 83.3% |
| | H12 | Proportion of patients with severe bacterial CAP given the appropriate IV/oral antibiotic dose according to the WHO AWaRe antibiotic book. | NA | NA | NA | 8 | A | 91.7% |
| | H13 | Proportion of patients with severe bacterial CAP given the appropriate IV/oral antibiotic duration according to the WHO AWaRe antibiotic book. | NA | NA | NA | 8 | A | 83.3% |
| | H14 | Proportion of patients presenting with severe* suspected bacterial community acquired pneumonia (CAP) given any Access antibiotic. | 8 | A | 81.3% | 7 | E | 75% |
| | H15 | Proportion of children presenting with severe* suspected bacterial community acquired pneumonia (CAP) given any Access antibiotic. | NA | NA | NA | 8 | A | 100% |
| | H16 | Proportion of patients presenting with severe* suspected bacterial community acquired pneumonia (CAP) given any Watch antibiotic. | 8 | A | 84.1% | 8 | A | 83.3% |

| Hospital Indication | Indicator list | Quality Indicator | Delphi Approp Median[1] | Delphi Approp Consensus[2] | Delphi Percent Agreement (n=107)[3] | RAND/UCLA Approp Median[1] | RAND/UCLA Approp Consensus[2] | RAND/UCLA Percent Agreement (n=12)[3] |
|---|---|---|---|---|---|---|---|---|
| **Respiratory Tract Infections (cont.)** | H17 | Proportion of adult patients presenting with severe* suspected bacterial community acquired pneumonia (CAP) given any Watch antibiotic. | NA | NA | NA | 8 | A | 83.3% |
| | H18 | Proportion of patients presenting with suspected bacterial CAP given the duration in days of IV antibiotics recommended in the WHO AWaRe antibiotic book. | 8 | A | 82.2% | 7 | A | 83.3% |
| | H19 | Proportion of patients presenting with suspected bacterial CAP given the duration in days of antibiotics recommended in the WHO AWaRe antibiotic book. | NA | NA | NA | 8 | A | 91.7% |
| | H20 | Proportion of patients presenting with suspected bacterial CAP prescribed the total daily dose of IV antibiotics in the WHO AWaRe antibiotic book. | NA | NA | NA | 6 | E | 66.7% |
| | H21 | Proportion of patients with all bacterial CAP given IV/oral Watch antibiotics. | 8 | A | 80.4% | NA | NA | NA |
| **Diarrhoea** | H22 | Proportion of patients with severe acute bloody diarrhoea given the appropriate antibiotic (drug choice and duration) according to the WHO AWaRe antibiotic book. | 8 | A | 87.9% | 8 | A | 83.3% |
| | H23 | Proportion of patients presenting with severe* acute bloody diarrhoea given the duration in days of IV antibiotics recommended in the WHO AWaRe antibiotic book. | 8 | A | 83.2% | 8 | A | 100% |
| | H24 | Proportion of patients presenting with severe* acute bloody diarrhoea prescribed the total daily dose of IV antibiotics in the WHO AWaRe antibiotic book. | NA | NA | NA | 7 | A | 100% |
| | H25 | Proportion of patients presenting with severe* acute bloody diarrhoea given Access antibiotics. | 8 | E | 76.6% | 8 | A | 91.7% |
| | H26 | Proportion of patients presenting with severe* acute bloody diarrhoea given Watch antibiotics. | 8 | A | 81.3% | 8 | A | 100% |

| Hospital Indication | Indicator list | Quality Indicator | Delphi Approp Median[1] | Delphi Approp Consensus[2] | Delphi Percent Agreement (n=107)[3] | RAND/UCLA Approp Median[1] | RAND/UCLA Approp Consensus[2] | RAND/UCLA Percent Agreement (n=12)[3] |
|---|---|---|---|---|---|---|---|---|
| **Intra-abdominal Infections** | H27 | Proportion of patients with mild-moderate intra-abdominal infections (IAIs) given the appropriate IV/oral antibiotic (drug choice and duration) according to the WHO AWaRe antibiotic book. | 8 | E | 79.4% | 7 | E | 75% |
| | H28 | Proportion of patients with mild-moderate intra-abdominal infections (IAIs) given the appropriate IV/oral antibiotic drug according to the WHO AWaRe antibiotic book. | NA | NA | NA | 7 | A | 83.3% |
| | H29 | Proportion of patients with mild-moderate intra-abdominal infections (IAIs) given the appropriate IV/oral antibiotic duration according to the WHO AWaRe antibiotic book. | NA | NA | NA | 8 | A | 91.7% |
| | H30 | Proportion of patients presenting with intra-abdominal infections (IAIs) (i.e. cholecystitis, cholangitis, acute appendicitis, acute diverticulitis) given IV antibiotics. | 8 | A | 83.2% | 8 | A | 83.3% |
| | H31 | Proportion of patients presenting with severe* intra-abdominal infections (IAIs) (i.e. cholecystitis, cholangitis, acute appendicitis, acute diverticulitis) given IV Access antibiotic. | 8 | E | 79.4% | 7 | A | 83.3% |
| | H32 | Proportion of patients presenting with severe* intra-abdominal infections (IAIs) (e.g. cholecystitis, cholangitis, acute appendicitis, acute diverticulitis) given IV Watch antibiotic. | 8 | A | 84.9% | 8 | A | 100% |
| | H33 | Proportion of patients with severe IAIs given the appropriate IV/oral antibiotic (drug choice and duration) according to the WHO AWaRe antibiotic book. | 8 | A | 84.1% | 7 | A | 91.7% |

| Hospital Indication | Indicator list | Quality Indicator | Delphi Approp Median[1] | Delphi Approp Consensus[2] | Delphi Percent Agreement (n=107)[3] | RAND/UCLA Approp Median[1] | RAND/UCLA Approp Consensus[2] | RAND/UCLA Percent Agreement (n=12)[3] |
|---|---|---|---|---|---|---|---|---|
| **Intra-abdominal Infections (cont.)** | H34 | Proportion of patients with severe IAIs given the appropriate IV/oral antibiotic according to the WHO AWaRe antibiotic book. | NA | NA | NA | 8 | A | 91.7% |
| | H35 | Proportion of patients with severe IAIs given the appropriate IV/oral antibiotic duration according to the WHO AWaRe antibiotic book. | NA | NA | NA | 7 | E | 75% |
| | H36 | Proportion of patients presenting with intra-abdominal infections (IAIs) (i.e. cholecystitis, cholangitis, acute appendicitis, acute diverticulitis) given the duration in days of IV antibiotics recommended in the WHO AWaRe antibiotic book. | 8 | A | 82.2% | 7 | A | 91.7% |
| | H37 | Proportion of patients presenting with intra-abdominal infections (IAIs) (i.e. cholecystitis, cholangitis, acute appendicitis, acute diverticulitis) prescribed the total daily dose of IV antibiotics in the WHO AWaRe antibiotic book. | NA | NA | NA | 7 | A | 100% |
| | H38 | Proportion of patients with all IAIs given IV/oral Watch antibiotics. | 8 | A | 81.3% | 8 | A | 91.7% |
| **Upper Urinary Tract Infection** | H39 | Proportion of patients with mild* upper urinary tract infection (UTIs) given the appropriate oral antibiotic (drug choice and duration) according to the WHO AWaRe antibiotic book. | 8 | A | 80.4% | 7 | A | 91.7% |
| | H40 | Proportion of patients with mild* upper urinary tract infection (UTIs) given the appropriate oral antibiotic according to the WHO AWaRe antibiotic book. | NA | NA | NA | 8 | E | 75% |
| | H41 | Proportion of patients with mild* upper urinary tract infection (UTIs) given the appropriate oral antibiotic duration according to the WHO AWaRe antibiotic book. | NA | NA | NA | 8 | A | 91.7% |

| Hospital Indication | Indicator list | Quality Indicator | Delphi Approp Median[1] | Delphi Approp Consensus[2] | Delphi Percent Agreement (n=107)[3] | RAND/UCLA Approp Median[1] | RAND/UCLA Approp Consensus[2] | RAND/UCLA Percent Agreement (n=12)[3] |
|---|---|---|---|---|---|---|---|---|
| **Upper Urinary Tract Infection (cont.)** | H42 | Proportion of patients with severe* upper UTIs given the appropriate IV antibiotics (drug choice and duration) according to the WHO AWaRe antibiotic book. | 8 | A | 86.8% | 7 | A | 100% |
| | H43 | Proportion of patients with severe* upper UTIs given the appropriate IV antibiotics according to the WHO AWaRe antibiotic book. | NA | NA | NA | 8 | A | 100% |
| | H44 | Proportion of patients with severe* upper UTIs given the appropriate IV antibiotics duration according to the WHO AWaRe antibiotic book. | NA | NA | NA | 8 | A | 91.7% |
| | H45 | Proportion of patients presenting with upper UTIs given the duration in days of IV antibiotics recommended in the WHO AWaRe antibiotic book. | 8 | A | 85.0% | 8 | A | 100% |
| | H46 | Proportion of patients presenting with upper UTIs prescribed the total daily dose of IV antibiotics in the WHO AWaRe antibiotic book. | NA | NA | NA | 7 | A | 100% |
| | H47 | Proportion of patients with all upper UTIs given IV/oral Watch antibiotics. | 8 | A | 80.2% | 7 | A | 83.3% |
| | H48 | Proportion of patients presenting with upper urinary tract infection (UTI) given IV antibiotics. | 8 | E | 76.6% | 8 | A | 83.3% |
| | H49 | Proportion of patients presenting with severe* upper urinary tract infection (UTI) given IV Access antibiotic. | 8 | A | 80.4% | 8 | A | 83.3% |
| | H50 | Proportion of patients presenting with severe* upper urinary tract infection (UTI) given IV Watch antibiotic. | 8 | A | 81.1% | 8 | A | 91.7% |
| **Skin/Soft Tissue Infections and Osteoarticular Infections** | H51 | Proportion of patients presenting with complicated skin and soft tissue infections (cSSTIs) (necrotizing fasciitis, pyomyositis) given the appropriate IV antibiotics according to the WHO AWaRe antibiotic book. | 8 | A | 84.1% | 8 | A | 91.7% |

| Hospital Indication | Indicator list | Quality Indicator | Delphi Approp Median[1] | Delphi Approp Consensus[2] | Delphi Percent Agreement (n=107)[3] | RAND/UCLA Approp Median[1] | RAND/UCLA Approp Consensus[2] | RAND/UCLA Percent Agreement (n=12)[3] |
|---|---|---|---|---|---|---|---|---|
| **Skin/Soft Tissue Infections and Osteoarticular Infections (cont.)** | H52 | Proportion of patients presenting with complicated skin and soft tissue infections (cSSTIs) (necrotizing fasciitis, pyomyositis) given the duration in days of IV antibiotics recommended in the WHO AWaRe antibiotic book. | 8 | E | 79.4% | 7 | E | 75% |
| | H53 | Proportion of patients presenting with complicated skin and soft tissue infections (cSSTIs) (necrotizing fasciitis, pyomyositis) prescribed the total daily dose of IV antibiotics in the WHO AWaRe antibiotic book. | NA | NA | NA | 7 | A | 83.3% |
| | H54 | Proportion of patients presenting with osteoarticular infections (acute bacterial osteomyelitis, septic arthritis) given IV Access antibiotic. | 8 | E | 78.5% | 7 | E | 75% |
| | H55 | Proportion of patients presenting with osteoarticular infections (acute bacterial osteomyelitis, septic arthritis) given IV Watch antibiotic. | 8 | E | 76.6% | 7 | E | 75% |
| | H56 | Proportion of patients presenting with osteoarticular infections (acute bacterial osteomyelitis, septic arthritis) given the duration in days of IV antibiotics recommended in the WHO AWaRe antibiotic book. | 8 | A | 81.3% | 7 | A | 83.3% |
| | H57 | Proportion of patients presenting with osteoarticular infections (acute bacterial osteomyelitis, septic arthritis) prescribed the total daily dose of IV antibiotics in the WHO AWaRe antibiotic book. | NA | NA | NA | 7 | A | 91.7% |
| **Surgical Prophylaxis** | H58 | Proportion of patients who received the appropriate IV surgical prophylaxis according to the WHO AWaRe antibiotic book for bowel surgery/clean or clean-contaminated procedure/contaminated procedure/urologic procedure. | 8 | A | 88.8% | 8 | A | 91.7% |

| Hospital Indication | Indicator list | Quality Indicator | Delphi Approp Median[1] | Delphi Approp Consensus[2] | Delphi Percent Agreement (n=107)[3] | RAND/UCLA Approp Median[1] | RAND/UCLA Approp Consensus[2] | RAND/UCLA Percent Agreement (n=12)[3] |
|---|---|---|---|---|---|---|---|---|
| **Surgical Prophylaxis (cont.)** | H59 | Proportion of patients given the duration in days of IV antibiotics recommended in the WHO AWaRe antibiotic book for bowel surgery/clean or clean-contaminated procedure/contaminated procedure/urologic procedure. | 8 | A | 83.2% | 8 | A | 100% |
| | H60 | Proportion of patients who received IV surgical prophylaxis at the appropriate timing according to the WHO AWaRe antibiotic book for bowel surgery/clean or clean-contaminated procedure/contaminated procedure/urologic procedure. | 8 | A | 90.6% | 8 | A | 100% |
| | H61 | Proportion of patients who received IV surgical prophylaxis at the appropriate timing according to the WHO AWaRe antibiotic book. | 8 | A | 90.7% | 8 | A | 100% |

[1] Panellist Median Rating on scale of 1-9
[2] Consensus Rating: A=Agreement, E=Equivocal, D=Disagreement, NA= Indicator not rated in round
[3] Percent of panellists that rated within the three-point scale of the median
*Severity criteria as per AWaRe guidance

#### Table S3: Appropriateness Ratings of General Model QIs Rated in the Delphi Technique and RAND/UCLA Method

| General Indication | Indicator list | Quality Indicator | Delphi Approp Median[1] | Delphi Approp Consensus[2] | Delphi Percent Agreement (n=107)[3] | Rand/UCLA Approp Median[1] | Rand/UCLA Approp Consensus[2] | Rand/UCLA Percent Agreement (n=12)[3] |
|---|---|---|---|---|---|---|---|---|
| **Patient Level** | G1 | The empiric treatment plan should be documented in the medical records and should include clinical diagnosis and indication for antibiotic treatment, dose and route of administration, intended duration of treatment, stop or review date, and IV to oral switch. | 8 | A | 93.5% | 8 | A | 100 |
| | G2 | Allergy status of the patient including timing, nature and severity of previous exposure/possible allergic reactions to antibiotics and the name/s of the antibiotic should be documented in the medical records. | 9 | A | 84.1% | 8 | A | 100 |
| | G3 | Antibiotics started for empiric therapy should be reviewed when any microbiological results from the patient become available and the decision to stop or change or continue the empirical therapy should be documented in the medical records. | 9 | A | 85.0% | 8 | A | 91.7 |
| | G4 | Antibiotics started for empiric therapy should be reviewed according to clinical response 48-72 hours after the start and the decision to stop or change or continue the empirical therapy should be documented in the medical records. | 8 | A | 94.4% | 8 | A | 91.7 |
| | G5 | Proportion of patients who are eligible for IV to oral switch, in whom intravenous route of administration is switched to the oral route at an appropriate time. | 8 | A | 88.8% | 8 | A | 83.3 |
| | G6 | Any toxicity/adverse reaction to an antibiotic (including type, duration of symptoms, time of onset since antibiotic administration) should be documented in medical records. | 8 | A | 88.8% | 8 | A | 91.7 |

| General Indication | Indicator list | Quality Indicator | Delphi Approp Median[1] | Delphi Approp Consensus[2] | Delphi Percent Agreement (n=107)[3] | Rand/UCLA Approp Median[1] | Rand/UCLA Approp Consensus[2] | Rand/UCLA Percent Agreement (n=12)[3] |
|---|---|---|---|---|---|---|---|---|
| **Patient Level (cont.)** | G7 | Blood cultures should be performed in all patients with severe bacterial infections (sepsis or septic shock, severe pneumonia, meningitis, severe IAIs, severe UTIs) ideally before starting antibiotics or as soon as possible if not sent before. | 9 | A | 82.3% | 8 | A | 100 |
| | G8 | Proportion of patients with suspected multi-drug resistant Gram-negative infections given Reserve antibiotics for empiric use according to the WHO AWaRe antibiotic book. | 8 | A | 87.9% | 7 | E | 66.7 |
| | G9 | Proportion of patients with suspected multi-drug resistant infections given Reserve antibiotics for empiric use according to the WHO AWaRe antibiotic book. | NA | NA | NA | 8 | A | 83.3 |
| | G10 | Proportion of patients with severe illness given Reserve antibiotics for empiric use according to the WHO AWaRe antibiotic book. | NA | NA | NA | 7 | E | 58.3 |
| | G11 | Proportion of patients with suspected multi-drug resistant Gram-positive infections given Reserve antibiotics for empiric use according to the WHO AWaRe antibiotic book. | 8 | A | 86.9% | 6 | E | 50 |
| **Population Level** | G12 | Use of oral Access and Watch antibiotics (split by AWaRe group) measured in DID (Defined daily doses per 1000 inhabitants per day) in Primary Care. | 8 | A | 90.7% | 8 | A | 83.3 |
| | G13 | Use of IV and oral Access and Watch and Reserve antibiotics (split by AWaRe group) measured in DDD (Defined daily doses) per 100 hospital admissions per day. | 8 | A | 85% | 8 | E | 75 |
| | G14 | Use of IV and oral Access and Watch and Reserve antibiotics (split by AWaRe group) measured in DDD (Defined daily doses) per 100 patient days (hospital occupied bed days). | NA | NA | NA | 8 | A | 91.7 |

| General Indication | Indicator list | Quality Indicator | Delphi Approp Median[1] | Delphi Approp Consensus[2] | Delphi Percent Agreement (n=107)[3] | Rand/UCLA Approp Median[1] | Rand/UCLA Approp Consensus[2] | Rand/UCLA Percent Agreement (n=12)[3] |
|---|---|---|---|---|---|---|---|---|
| **Population Level (cont.)** | G15 | At least 80% of total oral antibiotic use in primary care should be Access antibiotics. | 7 | A | 72.9% | 8 | E | 75 |
| | G16 | Percentage of total oral Access antibiotic use. | NA | NA | NA | 8 | A | 100 |
| | G17 | At least 70% of total country level antibiotic consumption should be Access antibiotics. | 7 | E | 70.1% | 8 | E | 75 |
| | G18 | Proportion of antibiotics given for respiratory infections during the high-prevalence season (e.g. winter-summer or wet-dry)<br>Compared to The proportion of antibiotics given for respiratory infections given during the low-prevalence season (e.g. summer-winter or dry-wet) | 7 | E | 62.6% | NA | NA | NA |
| | G19 | Percentage of total country level Access antibiotic use | NA | NA | NA | 8 | A | 100 |
| | G20 | Ratio of oral amoxicillin measured in DID (Defined daily doses per 1000 inhabitants per day) in primary care to all oral antibiotics in primary care measured in DID excluding amoxicillin (including amoxicillin-clavulanic acid) | 7 | E | 63.6% | 7 | A | 83.3 |
| | G21 | Ratio of oral "Not Recommended" antibiotic measured in DID (Defined daily doses per 1000 inhabitants per day) to all oral antibiotics measured in DID (J01) | 8 | A | 82.2% | 7 | E | 75 |
| | G22 | Number of primary care prescriptions >5 days for amoxicillin (J01CA04)<br>Over Total number of primary care antibiotic prescriptions for amoxicillin | 7 | E | 72.0% | NA | NA | NA |

[1] Panellist Median Rating on scale of 1-9
[2] Consensus Rating: A=Agreement, E=Equivocal, D=Disagreement, NA= Indicator not rated in round
[3] Percent of panellists that rated within the three-point scale of the median

#### Table S4: Feasibility Ratings of Primary Care model Quality Indicators Rated in the Delphi Technique and RAND/UCLA Panel

| Primary Care Indication | Indicator list | Quality Indicator | Delphi Feas Median[1] | Delphi Feas Consensus[2] | Delphi Feas Percent Agreement[3] | RAND/UCLA Feas Median[1] | RAND/UCLA Feas Consensus[2] | RAND/UCLA Feas Percent Agreement[3] |
|---|---|---|---|---|---|---|---|---|
| **Respiratory Tract Infections** | P1 | Proportion of all patients presenting with any acute respiratory tract infection (RTI) given an oral antibiotic. | 8 | E | 77.6% | 8 | E | 66.7% |
| | P2 | Proportion of all patients presenting with any acute respiratory tract infection (RTI) given oral amoxicillin. | 7 | E | 70.1% | 8 | E | 75% |
| | P3 | Proportion of all patients presenting with any acute respiratory tract infection (RTI) given any oral Access antibiotic (including amoxicillin). | 8 | E | 75.7% | 8 | E | 75% |
| | P4 | Proportion of all patients presenting with any acute respiratory tract infection (RTI) given any oral Watch antibiotic. | 8 | E | 72% | 7 | A | 91.7% |
| | P5 | Proportion of patients presenting with bronchitis given an oral antibiotic. | 7 | E | 69.8% | 7 | A | 83.3% |
| | P6 | Proportion of patients with any ear/sinus/throat infection (not pneumonia) given an oral antibiotic. | 8 | E | 75.7% | 8 | A | 83.3% |
| | P7 | Proportion of patients with any ear/sinus/throat infection (not pneumonia) at high risk* of severe complications given an oral antibiotic. | 7 | E | 69.2% | 8 | E | 66.7% |
| | P8 | Proportion of patients with any ear/sinus/throat infection (not pneumonia) at high risk* of severe complications given amoxicillin. | 7 | E | 69.8% | 7 | E | 75% |

| Primary Care Indication | Indicator list | Quality Indicator | Delphi Feas Median[1] | Delphi Feas Consensus[2] | Delphi Feas Percent Agreement[3] | RAND/UCLA Feas Median[1] | RAND/UCLA Feas Consensus[2] | RAND/UCLA Feas Percent Agreement[3] |
|---|---|---|---|---|---|---|---|---|
| **Respiratory Tract Infections (cont.)** | P9 | Proportion of patients with any ear/sinus/throat infection (not pneumonia) at high risk* of severe complications given any oral Access antibiotic (including amoxicillin). | 7 | E | 72% | 7 | A | 91.7% |
| | P10 | Proportion of patients at lower risk of a bacterial respiratory tract infection given an oral antibiotic. | 6 | E | 65.4% | NA | NA | 50% |
| | P11 | Proportion of patients with acute RTIs given the duration in days of oral antibiotics recommended in the WHO AWaRe antibiotic book | 7 | E | 71% | 6 | E | 50% |
| | P12 | Proportion of patients with acute RTIs prescribed the total daily dose of oral antibiotics recommended in the WHO AWaRe antibiotic book. | NA | NA | NA | 6 | E | 66.7% |
| | P13 | Proportion of patients with bacterial RTIs given oral Access or Watch antibiotics. | 7 | E | 73.8% | 6 | E | 41.7% |
| **Dental Infections** | P14 | Proportion of otherwise healthy adults presenting with dental infections given an oral antibiotic. | 7 | E | 65.4% | 6 | E | 41.7% |
| | P15 | Proportion of otherwise healthy adults presenting with severe* dental infections given an oral antibiotic. | 7 | E | 70.1% | 6 | E | 41.7% |
| | P16 | Proportion of patients presenting with dental infections at high risk* of severe complications given an oral antibiotic. | 7 | E | 70.1% | 5 | E | 58.3% |
| | P17 | Proportion of patients presenting with dental infections at high risk* of severe complications given amoxicillin. | 7 | E | 66.4% | 6 | E | 50% |

| Primary Care Indication | Indicator list | Quality Indicator | Delphi Feas Median[1] | Delphi Feas Consensus[2] | Delphi Feas Percent Agreement[3] | RAND/UCLA Feas Median[1] | RAND/UCLA Feas Consensus[2] | RAND/UCLA Feas Percent Agreement[3] |
|---|---|---|---|---|---|---|---|---|
| **Dental Infections (cont.)** | P18 | Proportion of patients presenting with dental infections at high risk* of severe complications given any oral Access antibiotic (including amoxicillin). | 7 | E | 68.2% | 6 | E | 50% |
| | P19 | Proportion of patients with lower risk (of bacterial infection) dental infections given oral antibiotics. | 6 | E | 59.8% | 6 | E | 25% |
| | P20 | Proportion of patients presenting with dental infections given the duration in days of oral antibiotics recommended in the WHO AWaRe antibiotic book. | 7 | E | 72.6% | 5 | E | 50% |
| | P21 | Proportion of patients with dental infections prescribed the total daily dose of oral antibiotic recommended in the WHO AWaRe antibiotic book. | NA | NA | NA | 6 | E | 58.3% |
| | P22 | Proportion of patients with dental infections given oral Watch antibiotics. | 7 | A | 80.4% | 7 | E | 75% |
| **Bacterial Eye Infection** | P23 | Proportion of patients presenting with bacterial eye infections given the duration in days of oral antibiotics recommended in the WHO AWaRe antibiotic book. | 7 | E | 68.2% | 4 | E | 41.7% |
| | P24 | Proportion of patients with bacterial eye infections prescribed the total daily dose of oral antibiotics recommended in the WHO AWaRe antibiotic book. | NA | NA | NA | 4 | E | 50% |
| | P25 | Proportion of patients with eye infections prescribed the total daily dose of oral antibiotics recommended in the WHO AWaRe antibiotic book | NA | NA | NA | 5 | E | 58.3% |
| | P26 | Proportion of patients presenting with eye infection given an oral antibiotic. | 7 | E | 67.3% | 8 | E | 75% |

| Primary Care Indication | Indicator list | Quality Indicator | Delphi Feas Median[1] | Delphi Feas Consensus[2] | Delphi Feas Percent Agreement[3] | RAND/UCLA Feas Median[1] | RAND/UCLA Feas Consensus[2] | RAND/UCLA Feas Percent Agreement[3] |
|---|---|---|---|---|---|---|---|---|
| **Bacterial Eye Infections (cont.)** | P27 | Proportion of patients presenting with eye infection at high risk* of severe complications given any oral Access antibiotic. | 7 | E | 65.4% | 6 | E | 75% |
| | P28 | Proportion of patients presenting with eye infection at high risk* of severe complications given any oral Watch antibiotic. | 7 | E | 62.6% | 6 | E | 75% |
| **Diarrhoea** | P29 | Proportion of all patients presenting with acute infectious non-bloody diarrhoea given oral antibiotics. | 8 | E | 77.6% | 7 | A | 83.3% |
| | P30 | Proportion of all patients presenting with high-risk* acute infectious non-bloody diarrhoea given any oral Access antibiotic. | 7 | E | 74.8% | 7 | E | 58.3% |
| | P31 | Proportion of all patients presenting with high-risk* acute infectious non-bloody diarrhoea given any oral Watch antibiotic. | 7 | E | 72.9% | 6 | E | 50% |
| | P32 | Proportion of otherwise healthy patients presenting with acute infectious non-bloody diarrhoea given an oral antibiotic. | 7 | E | 72% | 6 | E | 66.7% |
| | P33 | Proportion of patients presenting with high-risk* acute infectious non-bloody diarrhoea given the duration in days of oral antibiotics recommended in the WHO AWaRe antibiotic book. | 7 | E | 69.2% | 4 | E | 75% |
| | P34 | Proportion of patients presenting with high-risk* acute infectious non-bloody diarrhoea prescribed the total daily dose of oral antibiotics recommended in the WHO AWaRe antibiotic book. | NA | NA | NA | 5 | E | 66.7% |
| | P35 | Proportion of patients with acute bloody infectious diarrhoea given oral Access or Watch antibiotics. | 8 | E | 73.8% | 7 | E | 75% |

| Primary Care Indication | Indicator list | Quality Indicator | Delphi Feas Median[1] | Delphi Feas Consensus[2] | Delphi Feas Percent Agreement[3] | RAND/UCLA Feas Median[1] | RAND/UCLA Feas Consensus[2] | RAND/UCLA Feas Percent Agreement[3] |
|---|---|---|---|---|---|---|---|---|
| **Diarrhoea (cont.)** | P36 | Proportion of patients with acute bloody infectious diarrhoea given oral Access antibiotics. | NA | NA | NA | 7 | E | 75% |
| | P37 | Proportion of patients with acute bloody infectious diarrhoea given oral Watch antibiotics. | NA | NA | NA | 7 | E | 66.7% |
| **Skin/Soft Tissue Infections and Lymphadenitis** | P38 | Proportion of patients presenting with acute lymphadenitis given an oral antibiotic. | 7 | E | 70.1% | 6 | E | 66.7% |
| | P39 | Proportion of patients presenting with higher risk* (of bacterial infection) acute lymphadenitis given an oral antibiotic. | 7 | E | 65.1% | 6 | E | 75% |
| | P40 | Proportion of patients presenting with higher risk* (of bacterial infection) acute lymphadenitis given any oral Access antibiotic. | 7 | E | 65.4% | 6 | E | 66.7% |
| | P41 | Proportion of patients presenting with higher risk* (of bacterial infection) acute lymphadenitis given any oral Watch antibiotic. | 7 | E | 63.5% | 6 | E | 58.3% |
| | P42 | Proportion of patients with lower risk (of bacterial infection) acute lymphadenitis given oral antibiotics. | 6 | E | 59.8% | 5 | E | 75% |
| | P43 | Proportion of patients presenting with acute lymphadenitis given the duration in days of oral antibiotics recommended in the WHO AWaRe antibiotic book. | 7 | E | 64.5% | 5 | E | 58.3% |
| | P44 | Proportion of patients presenting with acute lymphadenitis prescribed the total daily dose of oral antibiotics recommended in the WHO AWaRe antibiotic book. | NA | NA | NA | 5 | E | 75% |

| Primary Care Indication | Indicator list | Quality Indicator | Delphi Feas Median[1] | Delphi Feas Consensus[2] | Delphi Feas Percent Agreement[3] | RAND/UCLA Feas Median[1] | RAND/UCLA Feas Consensus[2] | RAND/UCLA Feas Percent Agreement[3] |
|---|---|---|---|---|---|---|---|---|
| **Skin/Soft Tissue Infections and Lymphadenitis (cont.)** | P45 | Proportion of patients presenting with mild* skin/soft tissue infections (SSTIs) given an oral antibiotic. | 7 | E | 74.8% | 7 | A | 83.3% |
| | P46 | Proportion of patients presenting with mild* skin/soft tissue infections (SSTIs) given a topical antibiotic. | NA | NA | NA | 7 | A | 83.3% |
| | P47 | Proportion of patients presenting with mild* skin/soft tissue infections (SSTIs) given any oral Access antibiotic. | 7 | E | 71% | 6 | E | 75% |
| | P48 | Proportion of patients presenting with mild* skin/soft tissue infections (SSTIs) given any oral Watch antibiotic. | 7 | E | 70.1% | 6 | E | 75% |
| | P49 | Proportion of patients presenting with mild* skin/soft tissue infections (SSTIs) given the duration in days of oral antibiotics recommended in the WHO AWaRe antibiotic book. | 7 | E | 70.8% | 5 | E | 66.7% |
| | P50 | Proportion of patients presenting with mild* skin/soft tissue infections (SSTIs) prescribed the total daily dose of oral antibiotics recommended in the WHO AWaRe antibiotic book. | NA | NA | NA | 6 | E | 58.3% |

| Primary Care Indication | Indicator list | Quality Indicator | Delphi Feas Median[1] | Delphi Feas Consensus[2] | Delphi Feas Percent Agreement[3] | RAND/UCLA Feas Median[1] | RAND/UCLA Feas Consensus[2] | RAND/UCLA Feas Percent Agreement[3] |
|---|---|---|---|---|---|---|---|---|
| **Lower urinary tract infections** | P51 | Proportion of low-risk patients* with positive urine test (positive urine leucocytes/leucocyte esterase or positive urine culture), but no urinary tract infection (UTI) symptoms (e.g., no dysuria, no increased urinary urgency and frequency, no lower abdominal pain or discomfort or sometimes visible hematuria), given oral antibiotics. | 7 | E | 63.6% | 5 | E | 50% |
| | P52 | Proportion of patients presenting with lower urinary tract infection (UTI) given an oral antibiotic. | 8 | A | 81.3% | 7 | A | 83.3% |
| | P53 | Proportion of patients presenting with lower urinary tract infection (UTI) given any oral Access antibiotic. | 8 | A | 85% | 7 | A | 83.3% |
| | P54 | Proportion of patients presenting with lower urinary tract infection (UTI) given any oral Watch antibiotic. | 8 | A | 86% | 7 | E | 75% |
| | P55 | Proportion of patients presenting with lower urinary tract infections (UTIs) given the duration in days of oral antibiotics recommended in the WHO AWaRe antibiotic book. | 7 | E | 74.8% | 5 | E | 58.3% |
| | P56 | Proportion of patients presenting with lower urinary tract infection (UTI) prescribed the total daily dose of oral antibiotics recommended in the WHO AWaRe antibiotic book. | NA | NA | NA | 6 | E | 50% |
| | P57 | Proportion of patients with lower urinary tract infection (UTIs) given oral Watch antibiotics. | 8 | A | 83.2% | 7 | E | 75% |

[1] Panellist Median Rating on scale of 1-9
[2] Consensus Rating: A=Agreement, E=Equivocal, D=Disagreement, NA= Indicator not rated in round
[3] Percent of panellists that rated within the three-point scale of the median
*Risk criteria as per AWaRe guidance
*Severity criteria as per AWaRe guidance

#### Table S5: Feasibility Ratings of Hospital model Quality Indicators Rated in the Delphi Technique and RAND/UCLA Panel

| Hospital Indication | Indicator list | Quality Indicator | Delphi Feas Median[1] | Delphi Feas Consensus[2] | Delphi Feas Percent Agreement[3] | RAND/UCLA Feas Median[1] | RAND/UCLA Feas Consensus[2] | RAND/UCLA Feas Percent Agreement[3] |
|---|---|---|---|---|---|---|---|---|
| **Undifferentiated sepsis** | H1 | Proportion of patients presenting with clinical sepsis of unknown origin given the appropriate IV antibiotics according to the WHO AWaRe antibiotic book. | 8 | E | 72% | 8 | E | 75% |
| | H2 | Proportion of patients presenting with clinical sepsis of unknown origin given the duration in days of IV empiric antibiotics recommended in the WHO AWaRe antibiotic book. | 7 | E | 73.8% | 7 | A | 83.3% |
| | H3 | Proportion of patients presenting with clinical sepsis of unknown origin prescribed the total daily dose of IV empiric antibiotics recommended in the WHO AWaRe antibiotic book. | NA | NA | NA | 7 | E | 66.7% |
| **Meningitis** | H4 | Proportion of patients presenting with bacterial meningitis given the appropriate IV antibiotics according to the WHO AWaRe antibiotic book. | 8 | E | 79.4% | 8 | E | 75% |
| | H5 | Proportion of patients presenting with presumed bacterial meningitis given the duration* in days of IV antibiotics recommended in the WHO AWaRe antibiotic book. | 8 | E | 74.8% | 7 | E | 75% |
| | H6 | Proportion of patients presenting with presumed bacterial meningitis prescribed the total daily dose of IV antibiotics in the WHO AWaRe antibiotic book. | NA | NA | NA | 6 | E | 58.3% |

| Hospital Indication | Indicator list | Quality Indicator | Delphi Feas Median[1] | Delphi Feas Consensus[2] | Delphi Feas Percent Agreement[3] | RAND/UCLA Feas Median[1] | RAND/UCLA Feas Consensus[2] | RAND/UCLA Feas Percent Agreement[3] |
|---|---|---|---|---|---|---|---|---|
| **Respiratory Tract Infections** | H7 | Proportion of patients presenting with non-severe* community acquired pneumonia (CAP) given amoxicillin. | 8 | E | 75.7% | 7 | A | 100% |
| | H8 | Proportion of patients presenting with non-severe* community acquired pneumonia (CAP) given any Access antibiotic. | 8 | E | 76.6% | 7 | A | 100% |
| | H9 | Proportion of patients presenting with non-severe* community acquired pneumonia (CAP) given any Watch antibiotic. | 8 | E | 75.7% | 7 | A | 100% |
| | H10 | Proportion of patients with severe bacterial community acquired pneumonia (CAP) given the appropriate IV/oral antibiotic (drug choice and duration) according to the WHO AWaRe antibiotic book. | 8 | E | 78.5% | 6 | E | 75% |
| | H11 | Proportion of patients with severe bacterial CAP given the appropriate IV/oral antibiotic drug according to the WHO AWaRe antibiotic book. | NA | NA | NA | 7 | A | 83.3% |
| | H12 | Proportion of patients with severe bacterial CAP given the appropriate IV/oral antibiotic dose according to the WHO AWaRe antibiotic book. | NA | NA | NA | 7 | E | 66.7% |
| | H13 | Proportion of patients with severe bacterial CAP given the appropriate IV/oral antibiotic duration according to the WHO AWaRe antibiotic book. | NA | NA | NA | 6 | E | 58.3% |

| Hospital Indication | Indicator list | Quality Indicator | Delphi Feas Median[1] | Delphi Feas Consensus[2] | Delphi Feas Percent Agreement[3] | RAND/UCLA Feas Median[1] | RAND/UCLA Feas Consensus[2] | RAND/UCLA Feas Percent Agreement[3] |
|---|---|---|---|---|---|---|---|---|
| **Respiratory Tract Infections (cont.)** | H14 | Proportion of patients presenting with severe* suspected bacterial community acquired pneumonia (CAP) given any Access antibiotic. | 7 | E | 77.6% | 7 | A | 83.3% |
| | H15 | Proportion of children presenting with severe* suspected bacterial community acquired pneumonia (CAP) given any Access antibiotic. | NA | NA | NA | 8 | E | 75% |
| | H16 | Proportion of patients presenting with severe* suspected bacterial community acquired pneumonia (CAP) given any Watch antibiotic. | 7 | E | 79.4% | 7 | A | 91.7% |
| | H17 | Proportion of adult patients presenting with severe* suspected bacterial community acquired pneumonia (CAP) given any Watch antibiotic. | NA | NA | NA | 7 | E | 75% |
| | H18 | Proportion of patients presenting with suspected bacterial CAP given the duration in days of IV antibiotics recommended in the WHO AWaRe antibiotic book. | 8 | E | 73.8% | 7 | E | 75% |
| | H19 | Proportion of patients presenting with suspected bacterial CAP given the duration in days of antibiotics recommended in the WHO AWaRe antibiotic book. | NA | NA | NA | 7 | E | 58.3% |
| | H20 | Proportion of patients presenting with suspected bacterial CAP prescribed the total daily dose of IV antibiotics in the WHO AWaRe antibiotic book. | NA | NA | NA | 6 | E | 66.7% |
| | H21 | Proportion of patients with all bacterial CAP given IV/oral Watch antibiotics. | 8 | E | 78.5% | NA | NA | NA |

| Hospital Indication | Indicator list | Quality Indicator | Delphi Feas Median[1] | Delphi Feas Consensus[2] | Delphi Feas Percent Agreement[3] | RAND/UCLA Feas Median[1] | RAND/UCLA Feas Consensus[2] | RAND/UCLA Feas Percent Agreement[3] |
|---|---|---|---|---|---|---|---|---|
| Diarrhoea | H22 | Proportion of patients with severe acute bloody diarrhoea given the appropriate antibiotic (drug choice and duration) according to the WHO AWaRe antibiotic book. | 8 | E | 75.7% | 7 | A | 83.3% |
| | H23 | Proportion of patients presenting with severe* acute bloody diarrhoea given the duration in days of IV antibiotics recommended in the WHO AWaRe antibiotic book. | 7 | E | 70.1% | 7 | A | 83.3% |
| | H24 | Proportion of patients presenting with severe* acute bloody diarrhoea prescribed the total daily dose of IV antibiotics in the WHO AWaRe antibiotic book. | NA | NA | NA | 6 | E | 75% |
| | H25 | Proportion of patients presenting with severe* acute bloody diarrhoea given Access antibiotics. | 8 | E | 73.8% | 7 | A | 91.7% |
| | H26 | Proportion of patients presenting with severe* acute bloody diarrhoea given Watch antibiotics. | 8 | E | 72.9% | 7 | A | 83.3% |
| Intra-abdominal Infections | H27 | Proportion of patients with mild-moderate intra-abdominal infections (IAIs) given the appropriate IV/oral antibiotic (drug choice and duration) according to the WHO AWaRe antibiotic book. | 7 | E | 77.6% | 6 | E | 58.3% |
| | H28 | Proportion of patients with mild-moderate intra-abdominal infections (IAIs) given the appropriate IV/oral antibiotic drug according to the WHO AWaRe antibiotic book. | NA | NA | NA | 6 | E | 75% |

| Hospital Indication | Indicator list | Quality Indicator | Delphi Feas Median[1] | Delphi Feas Consensus[2] | Delphi Feas Percent Agreement[3] | RAND/UCLA Feas Median[1] | RAND/UCLA Feas Consensus[2] | RAND/UCLA Feas Percent Agreement[3] |
|---|---|---|---|---|---|---|---|---|
| **Intra-abdominal Infections (cont.)** | H29 | Proportion of patients with mild-moderate intra-abdominal infections (IAIs) given the appropriate IV/oral antibiotic duration according to the WHO AWaRe antibiotic book. | NA | NA | NA | 6 | E | 66.7% |
| | H30 | Proportion of patients presenting with intra-abdominal infections (IAIs) (i.e. cholecystitis, cholangitis, acute appendicitis, acute diverticulitis) given IV antibiotics. | 8 | E | 79.4% | 7 | A | 100% |
| | H31 | Proportion of patients presenting with severe* intra-abdominal infections (IAIs) (i.e. cholecystitis, cholangitis, acute appendicitis, acute diverticulitis) given IV Access antibiotic. | 8 | E | 72.9% | 7 | A | 100% |
| | H32 | Proportion of patients presenting with severe* intra-abdominal infections (IAIs) (e.g. cholecystitis, cholangitis, acute appendicitis, acute diverticulitis) given IV Watch antibiotic. | 8 | E | 76.4% | 7 | A | 100% |
| | H33 | Proportion of patients with severe IAIs given the appropriate IV/oral antibiotic (drug choice and duration) according to the WHO AWaRe antibiotic book. | 7 | E | 72.9% | 7 | A | 83.3% |
| | H34 | Proportion of patients with severe IAIs given the appropriate IV/oral antibiotic according to the WHO AWaRe antibiotic book. | NA | NA | NA | 7 | E | 66.7% |
| | H35 | Proportion of patients with severe IAIs given the appropriate IV/oral antibiotic duration according to the WHO AWaRe antibiotic book. | NA | NA | NA | 6 | E | 50% |

| Hospital Indication | Indicator list | Quality Indicator | Delphi Feas Median[1] | Delphi Feas Consensus[2] | Delphi Feas Percent Agreement[3] | RAND/UCLA Feas Median[1] | RAND/UCLA Feas Consensus[2] | RAND/UCLA Feas Percent Agreement[3] |
|---|---|---|---|---|---|---|---|---|
| **Intra-abdominal Infections (cont.)** | H36 | Proportion of patients presenting with intra-abdominal infections (IAIs) (i.e. cholecystitis, cholangitis, acute appendicitis, acute diverticulitis) given the duration in days of IV antibiotics recommended in the WHO AWaRe antibiotic book. | 7 | E | 70.1% | 7 | E | 66.7% |
| | H37 | Proportion of patients presenting with intra-abdominal infections (IAIs) (i.e. cholecystitis, cholangitis, acute appendicitis, acute diverticulitis) prescribed the total daily dose of IV antibiotics in the WHO AWaRe antibiotic book. | NA | NA | NA | 7 | E | 75% |
| | H38 | Proportion of patients with all IAIs given IV/oral Watch antibiotics. | 8 | E | 78.5% | 7 | A | 91.7% |
| **Upper Urinary Tract Infections** | H39 | Proportion of patients with mild* upper urinary tract infection (UTIs) given the appropriate oral antibiotic (drug choice and duration) according to the WHO AWaRe antibiotic book. | 7 | E | 73.8% | 7 | E | 75% |
| | H40 | Proportion of patients with mild* upper urinary tract infection (UTIs) given the appropriate oral antibiotic according to the WHO AWaRe antibiotic book.... | NA | NA | NA | 7 | A | 83.3% |
| | H41 | Proportion of patients with mild* upper urinary tract infection (UTIs) given the appropriate oral antibiotic duration according to the WHO AWaRe antibiotic book. | NA | NA | NA | 6 | E | 66.7% |
| | H42 | Proportion of patients with severe* upper UTIs given the appropriate IV antibiotics (drug choice and duration) according to the WHO AWaRe antibiotic book. | 8 | E | 75.5% | 7 | A | 91.7% |

| Hospital Indication | Indicator list | Quality Indicator | Delphi Feas Median[1] | Delphi Feas Consensus[2] | Delphi Feas Percent Agreement[3] | RAND/UCLA Feas Median[1] | RAND/UCLA Feas Consensus[2] | RAND/UCLA Feas Percent Agreement[3] |
|---|---|---|---|---|---|---|---|---|
| **Upper Urinary Tract Infections (cont.)** | H43 | Proportion of patients with severe* upper UTIs given the appropriate IV antibiotics according to the WHO AWaRe antibiotic book. | NA | NA | NA | 7 | A | 100% |
| | H44 | Proportion of patients with severe* upper UTIs given the appropriate IV antibiotics duration according to the WHO AWaRe antibiotic book. | NA | NA | NA | 6 | E | 58.3% |
| | H45 | Proportion of patients presenting with upper UTIs given the duration in days of IV antibiotics recommended in the WHO AWaRe antibiotic book. | 8 | E | 74.8% | 7 | E | 75% |
| | H46 | Proportion of patients presenting with upper UTIs prescribed the total daily dose of IV antibiotics in the WHO AWaRe antibiotic book. | NA | NA | NA | 7 | E | 75% |
| | H47 | Proportion of patients with all upper UTIs given IV/oral Watch antibiotics. | 8 | E | 79.2% | 7 | A | 83.3% |
| | H48 | Proportion of patients presenting with upper urinary tract infection (UTI) given IV antibiotics. | 8 | E | 79.4% | 7 | E | 75% |
| | H49 | Proportion of patients presenting with severe* upper urinary tract infection (UTI) given IV Access antibiotic. | 8 | A | 80.4% | 7 | A | 91.7% |
| | H50 | Proportion of patients presenting with severe* upper urinary tract infection (UTI) given IV Watch antibiotic. | 8 | A | 80.2% | 7 | A | 91.7% |
| **Skin/Soft Tissue and Osteoarticular Infections** | H51 | Proportion of patients presenting with complicated skin and soft tissue infections (cSSTIs) (necrotizing fasciitis, pyomyositis) given the appropriate IV antibiotics according to the WHO AWaRe antibiotic book. | 8 | E | 78.5% | 7 | A | 91.7% |

| Hospital Indication | Indicator list | Quality Indicator | Delphi Feas Median[1] | Delphi Feas Consensus[2] | Delphi Feas Percent Agreement[3] | RAND/UCLA Feas Median[1] | RAND/UCLA Feas Consensus[2] | RAND/UCLA Feas Percent Agreement[3] |
|---|---|---|---|---|---|---|---|---|
| **Skin/Soft Tissue Infections and Osteoarticular Infections (cont.)** | H52 | Proportion of patients presenting with complicated skin and soft tissue infections (cSSTIs) (necrotizing fasciitis, pyomyositis) given the duration in days of IV antibiotics recommended in the WHO AWaRe antibiotic book. | 7 | E | 72% | 6 | E | 75% |
| | H53 | Proportion of patients presenting with complicated skin and soft tissue infections (cSSTIs) (necrotizing fasciitis, pyomyositis) prescribed the total daily dose of IV antibiotics in the WHO AWaRe antibiotic book. | NA | NA | NA | 6 | E | 75% |
| | H54 | Proportion of patients presenting with osteoarticular infections (acute bacterial osteomyelitis, septic arthritis) given IV Access antibiotic. | 8 | E | 76.6% | 7 | A | 100% |
| | H55 | Proportion of patients presenting with osteoarticular infections (acute bacterial osteomyelitis, septic arthritis) given IV Watch antibiotic. | 7 | E | 77.6% | 7 | A | 100% |
| | H56 | Proportion of patients presenting with osteoarticular infections (acute bacterial osteomyelitis, septic arthritis) given the duration in days of IV antibiotics recommended in the WHO AWaRe antibiotic book. | 7 | E | 69.2% | 6 | E | 66.7% |
| | H57 | Proportion of patients presenting with osteoarticular infections (acute bacterial osteomyelitis, septic arthritis) prescribed the total daily dose of IV antibiotics in the WHO AWaRe antibiotic book. | NA | NA | NA | 6 | A | 91.7% |

| Hospital Indication | Indicator list | Quality Indicator | Delphi Feas Median[1] | Delphi Feas Consensus[2] | Delphi Feas Percent Agreement[3] | RAND/UCLA Feas Median[1] | RAND/UCLA Feas Consensus[2] | RAND/UCLA Feas Percent Agreement[3] |
|---|---|---|---|---|---|---|---|---|
| **Surgical prophylaxis** | H58 | Proportion of patients who received the appropriate IV surgical prophylaxis according to the WHO AWaRe antibiotic book for bowel surgery/clean or clean-contaminated procedure/contaminated procedure/urologic procedure. | 8 | A | 82.2% | 7 | A | 83.3% |
| | H59 | Proportion of patients given the duration in days of IV antibiotics recommended in the WHO AWaRe antibiotic book for bowel surgery/clean or clean-contaminated procedure/contaminated procedure/urologic procedure. | 8 | A | 82.3% | 7 | A | 91.7% |
| | H60 | Proportion of patients who received IV surgical prophylaxis at the appropriate timing according to the WHO AWaRe antibiotic book for bowel surgery/clean or clean-contaminated procedure/contaminated procedure/urologic procedure. | 8 | E | 75.7% | 7 | A | 83.3% |
| | H61 | Proportion of patients who received IV surgical prophylaxis at the appropriate timing according to the WHO AWaRe antibiotic book. | 8 | E | 78.5% | 7 | E | 75% |

[1] Panellist Median Rating on scale of 1-9
[2] Consensus Rating: A=Agreement, E=Equivocal, D=Disagreement, NA= Indicator not rated in round
[3] Percent of panellists that rated within the three-point scale of the median
*Severity criteria as per AWaRe guidance

#### Table S6: Feasibility Ratings of General model Quality Indicators Rated in the Delphi Technique and RAND/UCLA Panel

| General Indication | Indicator list | Quality Indicator | Delphi Feas Median[1] | Delphi Feas Consensus[2] | Delphi Feas Percent Agreement[3] | RAND/UCLA Feas Median[1] | RAND/UCLA Feas Consensus[2] | RAND/UCLA Feas Percent Agreement[3] |
|---|---|---|---|---|---|---|---|---|
| **Patient Level** | G1 | The empiric treatment plan should be documented in the medical records and should include clinical diagnosis and indication for antibiotic treatment, dose and route of administration, intended duration of treatment, stop or review date, and IV to oral switch. | 7 | E | 65.4% | 6 | E | 58.3% |
| | G2 | Allergy status of the patient including timing, nature and severity of previous exposure/possible allergic reactions to antibiotics and the name/s of the antibiotic should be documented in the medical records. | 8 | E | 79.5% | 6 | E | 58.3% |
| | G3 | Antibiotics started for empiric therapy should be reviewed when any microbiological results from the patient become available and the decision to stop or change or continue the empirical therapy should be documented in the medical records. | 8 | A | 86% | 6 | E | 41.7% |
| | G4 | Antibiotics started for empiric therapy should be reviewed according to clinical response 48-72 hours after the start and the decision to stop or change or continue the empirical therapy should be documented in the medical records. | 8 | A | 85% | 6 | E | 66.7% |

| General Indication | Indicator list | Quality Indicator | Delphi Feas Median[1] | Delphi Feas Consensus[2] | Delphi Feas Percent Agreement[3] | RAND/UCLA Feas Median[1] | RAND/UCLA Feas Consensus[2] | RAND/UCLA Feas Percent Agreement[3] |
|---|---|---|---|---|---|---|---|---|
| **Patient Level (cont.)** | G5 | Proportion of patients who are eligible for IV to oral switch, in whom intravenous route of administration is switched to the oral route at an appropriate time. | 7 | E | 72% | 6 | E | 58.3% |
| | G6 | Any toxicity/adverse reaction to an antibiotic (including type, duration of symptoms, time of onset since antibiotic administration) should be documented in medical records. | 8 | E | 74.8% | 5 | E | 58.3% |
| | G7 | Blood cultures should be performed in all patient with severe bacterial infections (sepsis or septic shock, severe pneumonia, meningitis, severe IAIs, severe UTIs) ideally before starting antibiotics or as soon as possible if not sent before. | 8 | A | 84.1% | 5 | E | 66.7% |
| | G8 | Proportion of patients with suspected multi-drug resistant Gram-negative infections given Reserve antibiotics for empiric use according to the WHO AWaRe antibiotic book. | 8 | E | 79.4% | 6 | E | 58.3% |
| | G9 | Proportion of patients with suspected multi-drug resistant infections given Reserve antibiotics for empiric use according to the WHO AWaRe antibiotic book. | NA | NA | NA | 6 | E | 58.3% |
| | G10 | Proportion of patients with severe illness given Reserve antibiotics for empiric use according to the WHO AWaRe antibiotic book. | NA | NA | NA | 6 | E | 50% |
| | G11 | Proportion of patients with suspected multi-drug resistant Gram-positive infections given Reserve antibiotics for empiric use according to the WHO AWaRe antibiotic book. | 8 | E | 78.5% | 6 | E | 58.3% |

| General Indication | Indicator list | Quality Indicator | Delphi Feas Median[1] | Delphi Feas Consensus[2] | Delphi Feas Percent Agreement[3] | RAND/UCLA Feas Median[1] | RAND/UCLA Feas Consensus[2] | RAND/UCLA Feas Percent Agreement[3] |
|---|---|---|---|---|---|---|---|---|
| **Population Level** | G12 | Use of oral Access and Watch antibiotics (split by AWaRe group) measured in DID (Defined daily doses per 1000 inhabitants per day) in Primary Care. | 7 | E | 72.9% | 6 | E | 66.7% |
| | G13 | Use of IV and oral Access and Watch and Reserve antibiotics (split by AWaRe group) measured in DDD (Defined daily doses) per 100 hospital admissions per day. | 8 | A | 85% | 7 | A | 83.3% |
| | G14 | Use of IV and oral Access and Watch and Reserve antibiotics (split by AWaRe group) measured in DDD (Defined daily doses) per 100 patient days (hospital occupied bed days) | NA | NA | NA | 7 | A | 83.3% |
| | G15 | At least 80% of total oral antibiotic use in primary care should be Access antibiotics. | 7 | E | 72.9% | 6 | E | 58.3% |
| | G16 | Percentage of total oral Access antibiotic use. | NA | NA | NA | 8 | E | 75% |
| | G17 | At least 70% of total country level antibiotic consumption should be Access antibiotics. | 7 | E | 70.1% | 7 | E | 66.7% |
| | G18 | Proportion of antibiotics given for respiratory infections during the high-prevalence season (e.g. winter-summer or wet-dry) compared to the proportion of antibiotics given for respiratory infections given during the low-prevalence season (e.g. summer-winter or dry-wet) | 7 | E | 62.6% | NA | NA | NA |
| | G19 | Percentage of total country level Access antibiotic use. | NA | NA | NA | 8 | E | 66.7% |

| General Indication | Indicator list | Quality Indicator | Delphi Feas Median[1] | Delphi Feas Consensus[2] | Delphi Feas Percent Agreement[3] | RAND/UCLA Feas Median[1] | RAND/UCLA Feas Consensus[2] | RAND/UCLA Feas Percent Agreement[3] |
|---|---|---|---|---|---|---|---|---|
| **Population Level (cont.)** | G20 | Ratio of oral amoxicillin measured in DID (Defined daily doses per 1000 inhabitants per day) in primary care to all oral antibiotics in primary care measured in DID excluding amoxicillin (including amoxicillin-clavulanic acid) | 7 | E | 66.4% | 6 | A | 83.3% |
| | G21 | Ratio of oral "Not Recommended" antibiotics measured in DID (Defined daily doses per 1000 inhabitants per day) to all oral antibiotics measured in DID (J01) | 7 | E | 72.9% | 6 | E | 75% |
| | G22 | Number of primary care prescriptions?>?5 days for amoxicillin (J01CA04)<br><br>over<br><br>Total number of primary care antibiotic prescriptions for amoxicillin | 7 | E | 70.1% | NA | NA | NA |

[1] Panellist Median Rating on scale of 1-9
[2] Consensus Rating: A=Agreement, E=Equivocal, D=Disagreement, NA= Indicator not rated in round
[3] Percent of panellists that rated within the three-point scale of the median

#### Table S7: Quality Indicators Rated Both Appropriate and Feasible with Agreement by the Delphi Panel

| | Indication | Indicator list | Quality Indicator | Approp Median[1] | Approp Consensus[2] | Approp Percent Agreement[3] | Feas Median[1] | Feas Consensus[2] | Feas Percent Agreement[3] |
|---|---|---|---|---|---|---|---|---|---|
| **Primary Care Indicators** | **Dental Infections** | P22 | Proportion of patients with dental infections given oral Watch antibiotics. | 8 | A | 84.1% | 7 | A | 80.4% |
| | **Urinary Tract Infections** | P53 | Proportion of patients presenting with lower urinary tract infection (UTI) given any oral Access antibiotic. | 8 | A | 86.0% | 8 | A | 85% |
| | | P54 | Proportion of patients presenting with lower urinary tract infection (UTI) given any oral Watch antibiotic. | 8 | A | 86.9% | 8 | A | 86% |
| | | P57 | Proportion of patients with lower urinary tract infection (UTIs) given oral Watch antibiotics. | 8 | A | 85.0% | 8 | A | 83.2% |
| **Hospital Indicators** | **Urinary Tract Infections** | H49 | Proportion of patients presenting with severe* upper urinary tract infection (UTI) given IV Access antibiotic. | 8 | A | 80.4% | 8 | A | 80.4% |
| | | H50 | Proportion of patients presenting with severe* upper urinary tract infection (UTI) given IV Watch antibiotic. | 8 | A | 81.1% | 8 | A | 80.2% |
| | **Surgical Prophylaxis** | H58 | Proportion of patients who received the appropriate IV surgical prophylaxis according to the WHO AWaRe antibiotic book for bowel surgery/clean or clean-contaminated procedure/contaminated procedure/urologic procedure. | 8 | A | 88.8% | 8 | A | 82.2% |

| | Indication | Indicator list | Quality Indicator | Approp Median[1] | Approp Consensus[2] | Approp Percent Agreement[3] | Feas Median[1] | Feas Consensus[2] | Feas Percent Agreement[3] |
|---|---|---|---|---|---|---|---|---|---|
| **Hospital Indicators (cont.)** | **Surgical Prophylaxis (cont.)** | H59 | Proportion of patients given the duration in days of IV antibiotics recommended in the WHO AWaRe antibiotic book for bowel surgery/clean or clean-contaminated procedure/contaminated procedure/urologic procedure. | 8 | A | 83.2% | 8 | A | 82.3% |
| **General Indicators** | **Patient Level** | G3 | Antibiotics started for empiric therapy should be reviewed when any microbiological results from the patient become available and the decision to stop or change or continue the empirical therapy should be documented in the medical records. | 9 | A | 85.0% | 8 | A | 86% |
| | | G4 | Antibiotics started for empiric therapy should be reviewed according to clinical response 48-72 hours after the start and the decision to stop or change or continue the empirical therapy should be documented in the medical records. | 8 | A | 94.4% | 8 | A | 85% |
| | | G7 | Blood cultures should be performed in all patient with severe bacterial infections (sepsis or septic shock, severe pneumonia, meningitis, severe IAIs, severe UTIs) ideally before starting antibiotics or as soon as possible if not sent before. | 9 | A | 82.3% | 8 | A | 84.1% |
| | **Population Level** | G13 | Use of IV and oral Access and Watch and Reserve antibiotics (split by AWaRe group) measured in DDD (Defined daily doses) per 100 hospital admissions per day. | 8 | A | 85% | 8 | A | 85% |

[1] Panellist Median Rating on scale of 1-9
[2] Consensus Rating: A=Agreement, E=Equivocal, D=Disagreement, NA= Indicator not rated in round
[3] Percent of panellists that rated within the three-point scale of the median
*Risk criteria as per AWaRe guidance
*Severity criteria as per AWaRe guidance

#### Table S8: Quality Indicators Rated Both Appropriate and Feasible with Agreement by the RAND/UCLA Panel

| | Indication | Indicator list | Quality Indicator | Approp Median[1] | Approp Consensus[2] | Approp Percent Agreement[3] | Feas Median[1] | Feas Consensus[2] | Feas Percent Agreement[3] |
|---|---|---|---|---|---|---|---|---|---|
| **Primary Care** | **Respiratory Tract Infections** | P4 | Proportion of all patients presenting with any acute respiratory tract infection (RTI) given any oral Watch antibiotic. | 8 | A | 91.7% | 7 | A | 91.7% |
| | | P6 | Proportion of patients with any ear/sinus/throat infection (not pneumonia) given an oral antibiotic. | 8 | A | 83.3% | 8 | A | 83.3% |
| | | P9 | Proportion of patients with any ear/sinus/throat infection (not pneumonia) at high risk* of severe complications given any oral Access antibiotic (including amoxicillin). | 7 | A | 91.7% | 7 | A | 91.7% |
| | **Skin/Soft Tissue Infections and Lymphadenitis** | P45 | Proportion of patients presenting with mild* skin/soft tissue infections (SSTIs) given an oral antibiotic. | 7 | A | 100% | 7 | A | 83.3% |
| | | P46 | Proportion of patients presenting with mild* skin/soft tissue infections (SSTIs) given a topical antibiotic. | 7 | A | 83.3% | 7 | A | 83.3% |
| | **Urinary Tract Infections** | P52 | Proportion of patients presenting with lower urinary tract infection (UTI) given an oral antibiotic. | 8 | A | 91.7% | 7 | A | 83.3% |
| | | P53 | Proportion of patients presenting with lower urinary tract infection (UTI) given any oral Access antibiotic. | 8 | A | 91.7% | 7 | A | 83.3% |
| **Hospital** | **Undifferentiated Sepsis** | H2 | Proportion of patients presenting with clinical sepsis of unknown origin given the duration in days of IV empiric antibiotics recommended in the WHO AWaRe antibiotic book. | 8 | A | 91.7% | 7 | A | 83.3% |

| | Indication | Indicator list | Quality Indicator | Approp Median[1] | Approp Consensus[2] | Approp Percent Agreement [3] | Feas Median[1] | Feas Consensus[2] | Feas Percent Agreement[3] |
|---|---|---|---|---|---|---|---|---|---|
| **Hospital (cont.)** | **Respiratory Tract Infections** | H7 | Proportion of patients presenting with non-severe* community acquired pneumonia (CAP) given amoxicillin. | 8 | A | 83.3% | 7 | A | 100% |
| | | H9 | Proportion of patients presenting with non-severe* community acquired pneumonia (CAP) given any Watch antibiotic. | 8 | A | 83.3% | 7 | A | 100% |
| | | H11 | Proportion of patients with severe bacterial CAP given the appropriate IV/oral antibiotic drug according to the WHO AWaRe antibiotic book. | 8 | A | 83.3% | 7 | A | 83.3% |
| | | H16 | Proportion of patients presenting with severe* suspected bacterial community acquired pneumonia (CAP) given any Watch antibiotic. | 8 | A | 83.3% | 7 | A | 91.7% |
| | **Diarrhoea** | H22 | Proportion of patients with severe acute bloody diarrhoea given the appropriate antibiotic (drug choice and duration) according to the WHO AWaRe antibiotic book. | 8 | A | 83.3% | 7 | A | 83.3% |
| | | H23 | Proportion of patients presenting with severe* acute bloody diarrhoea given the duration in days of IV antibiotics recommended in the WHO AWaRe antibiotic book. | 8 | A | 100% | 7 | A | 83.3% |
| | | H25 | Proportion of patients presenting with severe* acute bloody diarrhoea given Access antibiotics. | 8 | A | 91.7% | 7 | A | 91.7% |
| | | H26 | Proportion of patients presenting with severe* acute bloody diarrhoea given Watch antibiotics. | 8 | A | 100% | 7 | A | 83.3% |

| | Indication | Indicator list | Quality Indicator | Approp Median[1] | Approp Consensus[2] | Approp Percent Agreement[3] | Feas Median[1] | Feas Consensus[2] | Feas Percent Agreement[3] |
|---|---|---|---|---|---|---|---|---|---|
| **Hospital (cont.)** | **Intra-abdominal Infections** | H30 | Proportion of patients presenting with intra-abdominal infections (IAIs) (i.e. cholecystitis, cholangitis, acute appendicitis, acute diverticulitis) given IV antibiotics. | 8 | A | 83.3% | 7 | A | 100% |
| | | H31 | Proportion of patients presenting with severe* intra-abdominal infections (IAIs) (i.e. cholecystitis, cholangitis, acute appendicitis, acute diverticulitis) given IV Access antibiotic. | 7 | A | 83.3% | 7 | A | 100% |
| | | H32 | Proportion of patients presenting with severe* intra-abdominal infections (IAIs) (e.g. cholecystitis, cholangitis, acute appendicitis, acute diverticulitis) given IV Watch antibiotic. | 8 | A | 100% | 7 | A | 100% |
| | | H33 | Proportion of patients with severe IAIs given the appropriate IV/oral antibiotic (drug choice and duration) according to the WHO AWaRe antibiotic book. | 7 | A | 91.7% | 7 | A | 83.3% |
| | | H38 | Proportion of patients with all IAIs given IV/oral Watch antibiotics. | 8 | A | 91.7% | 7 | A | 91.7% |
| | **Upper Urinary Tract Infections** | H42 | Proportion of patients with severe* upper UTIs given the appropriate IV antibiotics (drug choice and duration) according to the WHO AWaRe antibiotic book. | 7 | A | 100% | 7 | A | 91.7% |
| | | H43 | Proportion of patients with severe* upper UTIs given the appropriate IV antibiotics according to the WHO AWaRe antibiotic book. | 8 | A | 100% | 7 | A | 100% |
| | | H47 | Proportion of patients with all upper UTIs given IV/oral Watch antibiotics. | 7 | A | 83.3% | 7 | A | 83.3% |

| | Indication | Indicator list | Quality Indicator | Approp Median[1] | Approp Consensus[2] | Approp Percent Agreement[3] | Feas Median[1] | Feas Consensus[2] | Feas Percent Agreement[3] |
|---|---|---|---|---|---|---|---|---|---|
| **Hospital (cont.)** | **Upper Urinary Tract Infections (cont.)** | H49 | Proportion of patients presenting with severe* upper urinary tract infection (UTI) given IV Access antibiotic. | 8 | A | 83.3% | 7 | A | 91.7% |
| | | H50 | Proportion of patients presenting with severe* upper urinary tract infection (UTI) given IV Watch antibiotic. | 8 | A | 91.7% | 7 | A | 91.7% |
| | **Skin/Soft Tissue and Osteoarticular Infections** | H51 | Proportion of patients presenting with complicated skin and soft tissue infections (cSSTIs) (necrotizing fasciitis, pyomyositis) given the appropriate IV antibiotics according to the WHO AWaRe antibiotic book. | 8 | A | 91.7% | 7 | A | 91.7% |
| | **Surgical Prophylaxis** | H58 | Proportion of patients who received the appropriate IV surgical prophylaxis according to the WHO AWaRe antibiotic book for bowel surgery/clean or clean-contaminated procedure/contaminated procedure/urologic procedure. | 8 | A | 91.7% | 7 | A | 83.3% |
| | | H59 | Proportion of patients given the duration in days of IV antibiotics recommended in the WHO AWaRe antibiotic book for bowel surgery/clean or clean-contaminated procedure/contaminated procedure/urologic procedure. | 8 | A | 100% | 7 | A | 91.7% |
| | | H60 | Proportion of patients who received IV surgical prophylaxis at the appropriate timing according to the WHO AWaRe antibiotic book for bowel surgery/clean or clean-contaminated procedure/contaminated procedure/urologic procedure. | 8 | A | 100% | 7 | A | 83.3% |

| | Indication | Indicator list | Quality Indicator | Approp Median[1] | Approp Consensus[2] | Approp Percent Agreement[3] | Feas Median[1] | Feas Consensus[2] | Feas Percent Agreement[3] |
|---|---|---|---|---|---|---|---|---|---|
| **General** | **Population Level** | G14 | Use of IV and oral Access and Watch and Reserve antibiotics (split by AWaRe group) measured in DDD (Defined daily doses) per 100 patient days (hospital occupied bed days). | 8 | A | 91.7% | 7 | A | 83.3% |

[1] Panellist Median Rating on scale of 1-9
[2] Consensus Rating: A=Agreement, E=Equivocal, D=Disagreement, NA= Indicator not rated in round
[3] Percent of panellists that rated within the three-point scale of the median
*Risk criteria as per AWaRe guidance
*Severity criteria as per AWaRe guidance

#### 4. Priority and Supporting Indicators

##### Table S9: Primary Care Quality Indicators Prioritisation Process: Indicators reviewed and selected as priority and supporting indicators.

| Primary Care Indication | Indicator Number | Original Quality Indicator | Decisions and Rationale | Prioritised Quality Indicator |
|---|---|---|---|---|
| Respiratory Tract Infection | P1 | Proportion of all patients presenting with any acute respiratory tract infection (RTI) given an oral antibiotic. | Not Prioritised – negative indicator, does not align with WHO AWaRe book recommendations for this condition | |
| | P2 | Proportion of all patients presenting with any acute respiratory tract infection (RTI) given oral amoxicillin. | Prioritised – positive indicator, WHO recommended antibiotic choice | Proportion of all patients presenting with any acute respiratory tract infection (RTI) given oral amoxicillin. |
| | P3 | Proportion of all patients presenting with any acute respiratory tract infection (RTI) given any oral Access antibiotic (including amoxicillin). | Prioritised – positive indicator, WHO recommended antibiotic choice | Proportion of all patients presenting with any acute respiratory tract infection (RTI) given any oral Access antibiotic (including amoxicillin). |
| | P4 | Proportion of all patients presenting with any acute respiratory tract infection (RTI) given any oral Watch antibiotic. | Not Prioritised – negative indicator, does not align with WHO AWaRe book recommendations for this condition | |
| | P5 | Proportion of patients presenting with bronchitis given an oral antibiotic. | Prioritised – positive indicator for WHO recommendation of "No Antibiotic Care" , common overuse of antibiotics for this condition | Proportion of patients presenting with bronchitis given an oral antibiotic. |
| | P6 | Proportion of patients with any ear/sinus/throat infection (not pneumonia) given an oral antibiotic. | Not Prioritised – negative indicator, does not align with WHO AWaRe book recommendations for this condition | |

| **Primary Care Indication** | **Indicator Number** | **Original Quality Indicator** | **Decisions and Rationale** | **Prioritised Quality Indicator** |
|---|---|---|---|---|
| **Respiratory Tract Infection (cont.)** | P7 | Proportion of patients with any ear/sinus/throat infection (not pneumonia) at high risk* of severe complications given an oral antibiotic. | Not Prioritised – duplicate measurement of P8 | |
| | P8 | Proportion of patients with any ear/sinus/throat infection (not pneumonia) at high risk* of severe complications given amoxicillin. | Prioritised – positive indicator, WHO recommended antibiotic choice | Proportion of patients with any ear/sinus/throat infection (not pneumonia) at high risk* of severe complications given amoxicillin. |
| | P9 | Proportion of patients with any ear/sinus/throat infection (not pneumonia) at high risk* of severe complications given any oral Access antibiotic (including amoxicillin). | Not Prioritised – duplicate measurement of P8 | |
| | P10 | Proportion of patients at lower risk of a bacterial respiratory tract infection given an oral antibiotic. | Not Prioritised – negative indicator, does not align with WHO AWaRe book recommendations for this condition | |
| | P11 | Proportion of patients with acute RTIs given the duration in days of oral antibiotics recommended in the WHO AWaRe antibiotic book | Prioritised Supporting Indicator – positive indicator, WHO recommended antibiotic duration | *Proportion of patients with acute RTIs given the duration in days of oral antibiotics recommended in the WHO AWaRe antibiotic book* |
| | P12 | Proportion of patients with acute RTIs prescribed the total daily dose of oral antibiotics recommended in the WHO AWaRe antibiotic book. | Prioritised Supporting Indicator – positive indicator, WHO recommended antibiotic dose | *Proportion of patients with acute RTIs prescribed the total daily dose of oral antibiotics recommended in the WHO AWaRe antibiotic book.* |
| | P13 | Proportion of patients with bacterial RTIs given oral Access or Watch antibiotics. | Not Prioritised – duplicate measurement of P3 | |

| Primary Care Indication | Indicator Number | Original Quality Indicator | Decisions and Rationale | Prioritised Quality Indicator |
|---|---|---|---|---|
| **Dental Infection** | P14 | Proportion of otherwise healthy adults presenting with dental infections given an oral antibiotic. | Not Prioritised – negative indicator, does not align with WHO AWaRe book recommendations for this condition | |
| | P15 | Proportion of otherwise healthy adults presenting with severe* dental infections given an oral antibiotic. | Not Prioritised – duplicate measurement of P17 | |
| | P16 | Proportion of patients presenting with dental infections at high risk* of severe complications given an oral antibiotic. | Not Prioritised – duplicate measurement of P15 | |
| | P17 | Proportion of patients presenting with dental infections at high risk* of severe complications given amoxicillin. | Prioritised – positive indicator, WHO recommended antibiotic choice | Proportion of patients presenting with dental infections at higher risk of complications* and severe symptoms* given amoxicillin. |
| | P18 | Proportion of patients presenting with dental infections at high risk* of severe complications given any oral Access antibiotic (including amoxicillin). | Not Prioritised – duplicate measurement of P17 | |
| | P19 | Proportion of patients with lower risk (of bacterial infection) dental infections given oral antibiotics. | Not Prioritised – negative indicator, does not align with WHO AWaRe book recommendations for this condition | |
| | P20 | Proportion of patients presenting with dental infections given the duration in days of oral antibiotics recommended in the WHO AWaRe antibiotic book. | Prioritised Supporting Indicator – positive indicator, WHO recommended antibiotic duration | *Proportion of patients presenting with dental infections given the duration in days of oral antibiotics recommended in the WHO AWaRe antibiotic book.* |
| | P21 | Proportion of patients with dental infections prescribed the total daily dose of oral antibiotic recommended in the WHO AWaRe antibiotic book. | Prioritised Supporting Indicator – positive indicator, WHO recommended antibiotic dose | *Proportion of patients with dental infections prescribed the total daily dose of oral antibiotic recommended in the WHO AWaRe antibiotic book.* |
| | P22 | Proportion of patients with dental infections given oral Watch antibiotics. | Not Prioritised – negative indicator, does not align with WHO AWaRe book recommendations for this condition | |

| Primary Care Indication | Indicator Number | Original Quality Indicator | Decisions and Rationale | Prioritised Quality Indicator |
|---|---|---|---|---|
| **Bacterial Eye Infection** | P23 | Proportion of patients presenting with bacterial eye infections given the duration in days of oral antibiotics recommended in the WHO AWaRe antibiotic book. | Not Prioritised – negative indicator, does not align with WHO AWaRe book recommendations for this condition | |
| | P24 | Proportion of patients with bacterial eye infections prescribed the total daily dose of oral antibiotics recommended in the WHO AWaRe antibiotic book. | Not Prioritised – negative indicator, does not align with WHO AWaRe book recommendations for this condition | |
| | P25 | Proportion of patients with eye infections prescribed the total daily dose of oral antibiotics recommended in the WHO AWaRe antibiotic book | Not Prioritised – duplicate measurement of P24 | |
| | P26 | Proportion of patients presenting with eye infection given an oral antibiotic. | Not Prioritised – negative indicator, does not align with WHO AWaRe book recommendations for this condition | |
| | P27 | Proportion of patients presenting with eye infection at high risk* of severe complications given any oral Access antibiotic. | Not Prioritised – negative indicator, does not align with WHO AWaRe book recommendations for this condition | |
| | P28 | Proportion of patients presenting with eye infection at high risk* of severe complications given any oral Watch antibiotic. | Not Prioritised – negative indicator, does not align with WHO AWaRe book recommendations for this condition | |
| **Diarrhoea** | P29 | Proportion of all patients presenting with acute infectious non-bloody diarrhoea given oral antibiotics. | Prioritised – positive indicator for WHO recommendation of “No Antibiotic Care” , common overuse of antibiotics for this condition | |

| Primary Care Indication | Indicator Number | Original Quality Indicator | Decisions and Rationale | Prioritised Quality Indicator |
|---|---|---|---|---|
| **Diarrhoea (cont.)** | P30 | Proportion of all patients presenting with high-risk* acute infectious non-bloody diarrhoea given any oral Access antibiotic. | Not Prioritised – duplicate measurement of P29 | |
| | P31 | Proportion of all patients presenting with high-risk* acute infectious non-bloody diarrhoea given any oral Watch antibiotic. | Not Prioritised – negative indicator, does not align with WHO AWaRe book recommendations for this condition | |
| | P32 | Proportion of otherwise healthy patients presenting with acute infectious non-bloody diarrhoea given an oral antibiotic. | Not Prioritised – duplicate measurement of P29 | |
| | P33 | Proportion of patients presenting with high-risk* acute infectious non-bloody diarrhoea given the duration in days of oral antibiotics recommended in the WHO AWaRe antibiotic book. | Not Prioritised – negative indicator, does not align with WHO AWaRe book recommendations for this condition | |
| | P34 | Proportion of patients presenting with high-risk* acute infectious non-bloody diarrhoea prescribed the total daily dose of oral antibiotics recommended in the WHO AWaRe antibiotic book. | Not Prioritised – negative indicator, does not align with WHO AWaRe book recommendations for this condition | |
| | P35 | Proportion of patients with acute bloody infectious diarrhoea given oral Access or Watch antibiotics. | Not Prioritised – duplicate measurement of P37 | |
| | P36 | Proportion of patients with acute bloody infectious diarrhoea given oral Access antibiotics. | Not Prioritised – negative indicator, does not align with WHO AWaRe book recommendations for this condition | |
| | P37 | Proportion of patients with acute bloody infectious diarrhoea given oral Watch antibiotics. | Prioritised – positive indicator, WHO recommended antibiotic choice | Proportion of patients with acute bloody infectious diarrhoea given oral Watch antibiotics. |

| Primary Care Indication | Indicator Number | Original Quality Indicator | Decisions and Rationale | Prioritised Quality Indicator |
|---|---|---|---|---|
| **Skin/soft Tissue Infection and Lymphadenitis** | P38 | Proportion of patients presenting with acute lymphadenitis given an oral antibiotic. | Not Prioritised – rare/low burden of lymphadenitis in healthy adults[5] | |
| | P39 | Proportion of patients presenting with higher risk* (of bacterial infection) acute lymphadenitis given an oral antibiotic. | Not Prioritised – rare/low burden of lymphadenitis in healthy adults[5] | |
| | P40 | Proportion of patients presenting with higher risk* (of bacterial infection) acute lymphadenitis given any oral Access antibiotic. | Not Prioritised – rare/low burden of lymphadenitis in healthy adults[5] | |
| | P41 | Proportion of patients presenting with higher risk* (of bacterial infection) acute lymphadenitis given any oral Watch antibiotic. | Not Prioritised – rare/low burden of lymphadenitis in healthy adults[5] | |
| | P42 | Proportion of patients with lower risk (of bacterial infection) acute lymphadenitis given oral antibiotics. | Not Prioritised – rare/low burden of lymphadenitis in healthy adults[5] | |
| | P43 | Proportion of patients presenting with acute lymphadenitis given the duration in days of oral antibiotics recommended in the WHO AWaRe antibiotic book. | Not Prioritised – rare/low burden of lymphadenitis in healthy adults[5] | |
| | P44 | Proportion of patients presenting with acute lymphadenitis prescribed the total daily dose of oral antibiotics recommended in the WHO AWaRe antibiotic book. | Not Prioritised – rare/low burden of lymphadenitis in healthy adults[5] | |
| | P45 | Proportion of patients presenting with mild* skin/soft tissue infections (SSTIs) given an oral antibiotic. | Not Prioritised – duplicate measurement of P47 | |
| | P46 | Proportion of patients presenting with mild* skin/soft tissue infections (SSTIs) given a topical antibiotic. | Prioritised – positive indicator, WHO recommended antibiotic choice | Proportion of patients presenting with mild* skin/soft tissue infections (SSTIs) given a topical antibiotic. |
| | P47 | Proportion of patients presenting with mild* skin/soft tissue infections (SSTIs) given any oral Access antibiotic. | Prioritised – positive indicator, WHO recommended antibiotic choice | Proportion of patients presenting with mild* skin/soft tissue infections (SSTIs) given any oral Access antibiotic. |

| Primary Care Indication | Indicator Number | Original Quality Indicator | Decisions and Rationale | Prioritised Quality Indicator |
|---|---|---|---|---|
| **Skin/soft Tissue Infection and Lymphadenitis (cont.)** | P48 | Proportion of patients presenting with mild* skin/soft tissue infections (SSTIs) given any oral Watch antibiotic. | Not Prioritised – negative indicator, does not align with WHO AWaRe book recommendations for this condition | |
| | P49 | Proportion of patients presenting with mild* skin/soft tissue infections (SSTIs) given the duration in days of oral antibiotics recommended in the WHO AWaRe antibiotic book. | Prioritised Supporting Indicator – positive indicator, WHO recommended antibiotic duration | *Proportion of patients presenting with mild* skin/soft tissue infections (SSTIs) given the duration in days of oral antibiotics recommended in the WHO AWaRe antibiotic book.* |
| | P50 | Proportion of patients presenting with mild* skin/soft tissue infections (SSTIs) prescribed the total daily dose of oral antibiotics recommended in the WHO AWaRe antibiotic book. | Prioritised Supporting Indicator – positive indicator, WHO recommended antibiotic dose | *Proportion of patients presenting with mild* skin/soft tissue infections (SSTIs) prescribed the total daily dose of oral antibiotics recommended in the WHO AWaRe antibiotic book.* |
| **Lower Urinary Tract Infection** | P51 | Proportion of low-risk patients* with positive urine test (positive urine leucocytes/leucocyte esterase or positive urine culture), but no urinary tract infection (UTI) symptoms (e.g., no dysuria, no increased urinary urgency and frequency, no lower abdominal pain or discomfort or sometimes visible haematuria), given oral antibiotics. | Not Prioritised – Diagnostic uncertainty in measuring this indicator | |
| | P52 | Proportion of patients presenting with lower urinary tract infection (UTI) given an oral antibiotic. | Not Prioritised – duplicate measurement of P53 | |
| | P53 | Proportion of patients presenting with lower urinary tract infection (UTI) given any oral Access antibiotic. | Prioritised – positive indicator, WHO recommended antibiotic choice | Proportion of patients presenting with lower urinary tract infection (UTI) given any oral Access antibiotic. |
| | P54 | Proportion of patients presenting with lower urinary tract infection (UTI) given any oral Watch antibiotic. | Not Prioritised – negative indicator, does not align with WHO AWaRe book recommendations for this condition | |

| Primary Care Indication | Indicator Number | Original Quality Indicator | Decisions and Rationale | Prioritised Quality Indicator |
|---|---|---|---|---|
| **Lower Urinary Tract Infection (cont.)** | P55 | Proportion of patients presenting with lower urinary tract infections (UTIs) given the duration in days of oral antibiotics recommended in the WHO AWaRe antibiotic book. | Prioritised Supporting Indicator – positive indicator, WHO recommended antibiotic duration | *Proportion of patients presenting with lower urinary tract infections (UTIs) given the duration in days of oral antibiotics recommended in the WHO AWaRe antibiotic book.* |
| | P56 | Proportion of patients presenting with lower urinary tract infection (UTI) prescribed the total daily dose of oral antibiotics recommended in the WHO AWaRe antibiotic book. | Prioritised Supporting Indicator – positive indicator, WHO recommended antibiotic dose | *Proportion of patients presenting with lower urinary tract infection (UTI) prescribed the total daily dose of oral antibiotics recommended in the WHO AWaRe antibiotic book.* |
| | P57 | Proportion of patients with lower urinary tract infection (UTIs) given oral Watch antibiotics. | Not Prioritised – duplicate measurement of P54 | |

*Red highlighted words reflect changes made during tidying review process
*Italicised priority indicators are supporting indicators measuring duration and dose of antibiotic

Table S10: Hospital Quality Indicators Prioritisation Process: Indicators reviewed and selected as priority and supporting indicators.

| Hospital Indication | Indicator Number | Original Quality Indicator | Discussion edits | Prioritised Quality Indicator |
|---|---|---|---|---|
| **Undifferentiated Sepsis** | H1 | Proportion of patients presenting with clinical sepsis of unknown origin given the appropriate IV antibiotics according to the WHO AWaRe antibiotic book. | Prioritised – positive indicator, WHO recommended antibiotic choice | Proportion of patients presenting with clinical sepsis of unknown origin given the appropriate IV antibiotics according to the WHO AWaRe antibiotic book. |
| | H2 | Proportion of patients presenting with clinical sepsis of unknown origin given the duration in days of IV empiric antibiotics recommended in the WHO AWaRe antibiotic book. | Prioritised Supporting Indicator – positive indicator, WHO recommended antibiotic duration | *Proportion of patients presenting with clinical sepsis of unknown origin given the duration in days of IV empiric antibiotics recommended in the WHO AWaRe antibiotic book.* |
| | H3 | Proportion of patients presenting with clinical sepsis of unknown origin prescribed the total daily dose of IV empiric antibiotics recommended in the WHO AWaRe antibiotic book. | Prioritised Supporting Indicator – positive indicator, WHO recommended antibiotic dose | *Proportion of patients presenting with clinical sepsis of unknown origin prescribed the total daily dose of IV empiric antibiotics recommended in the WHO AWaRe antibiotic book.* |
| **Meningitis** | H4 | Proportion of patients presenting with suspected bacterial meningitis given the appropriate IV antibiotics according to the WHO AWaRe antibiotic book. | During Tidying Process– Add in word "suspected" as it cannot be empirically confirmed bacterial meningitis<br>Prioritised – positive indicator, WHO recommended antibiotic choice | Proportion of patients presenting with bacterial meningitis given the appropriate IV antibiotics according to the WHO AWaRe antibiotic book. |
| | H5 | Proportion of patients presenting with suspected bacterial meningitis given the duration in days of IV antibiotics recommended in the WHO AWaRe antibiotic book. | During Tidying Process– Add in word "suspected" as it cannot be empirically confirmed bacterial meningitis<br>Prioritised Supporting Indicator – positive indicator, WHO recommended antibiotic duration | *Proportion of patients presenting with presumed bacterial meningitis given the duration in days of IV antibiotics recommended in the WHO AWaRe antibiotic book.* |

| Hospital Indication | Indicator Number | Original Quality Indicator | Discussion edits | Prioritised Quality Indicator |
|---|---|---|---|---|
| **Meningitis (cont.)** | H6 | Proportion of patients presenting with suspected bacterial meningitis prescribed the total daily dose of IV antibiotics in the WHO AWaRe antibiotic book. | During Tidying Process– Add in word "suspected" as it cannot be empirically confirmed bacterial meningitis<br>Prioritised Supporting Indicator – positive indicator, WHO recommended antibiotic dose | *Proportion of patients presenting with presumed bacterial meningitis prescribed the total daily dose of IV antibiotics in the WHO AWaRe antibiotic book.* |
| **Respiratory Tract Infection** | H7 | Proportion of patients presenting with mild/moderate community acquired pneumonia (CAP) given amoxicillin. | Not Prioritised – duplicate measurement of H8 | |
| | H8 | Proportion of patients presenting with mild/moderate community acquired pneumonia (CAP) given any Access antibiotic. | During Tidying Process– Change "non-severe" to "mild/moderate" as that is in line with language in WHO AWaRe book<br>Prioritised – positive indicator, WHO recommended antibiotic choice | Proportion of patients presenting with non-severe* community acquired pneumonia (CAP) given any Access antibiotic. |
| | H9 | Proportion of patients presenting with mild/moderate community acquired pneumonia (CAP) given any Watch antibiotic. | Not Prioritised – negative indicator, does not align with WHO AWaRe book recommendations for this condition | |
| | H10 | Proportion of patients with severe bacterial community acquired pneumonia (CAP) given the appropriate IV/oral antibiotic (drug choice and duration) according to the WHO AWaRe antibiotic book. | Not Prioritised – duplicate measurement of H11 & H13 | |
| | H11 | Proportion of patients with severe bacterial CAP given the appropriate IV/oral antibiotic drug according to the WHO AWaRe antibiotic book. | Not Prioritised – Diagnostic uncertainty in measuring this indicator | |

| Hospital Indication | Indicator Number | Original Quality Indicator | Discussion edits | Prioritised Quality Indicator |
|---|---|---|---|---|
| **Respiratory Tract Infection (cont.)** | H12 | Proportion of patients with severe bacterial CAP given the appropriate IV/oral antibiotic total daily dose according to the WHO AWaRe antibiotic book. | Not Prioritised – Diagnostic uncertainty in measuring this indicator | |
| | H13 | Proportion of patients with severe bacterial CAP given the appropriate IV/oral antibiotic duration in days according to the WHO AWaRe antibiotic book. | Not Prioritised – Diagnostic uncertainty in measuring this indicator | |
| | H14 | Proportion of patients presenting with severe* suspected bacterial community acquired pneumonia (CAP) given any Access antibiotic. | Not Prioritised – negative indicator, does not align with WHO AWaRe book recommendations for this condition | |
| | H15 | Proportion of children presenting with severe* suspected bacterial community acquired pneumonia (CAP) given any Access antibiotic. | Not Prioritised – negative indicator, does not align with WHO AWaRe book recommendations for this condition | |
| | H16 | Proportion of patients presenting with severe* suspected bacterial community acquired pneumonia (CAP) given any Watch antibiotic. | Prioritised – positive indicator, WHO recommended antibiotic choice | Proportion of patients presenting with severe* suspected bacterial community acquired pneumonia (CAP) given any Watch antibiotic. |
| | H17 | Proportion of adult patients presenting with severe* suspected bacterial community acquired pneumonia (CAP) given any Watch antibiotic. | Not Prioritised – duplicate measurement of H16 | |
| | H18 | Proportion of patients presenting with suspected bacterial CAP given the duration in days of IV antibiotics recommended in the WHO AWaRe antibiotic book. | Not Prioritised – duplicate measurement of H19 | |

| Hospital Indication | Indicator Number | Original Quality Indicator | Discussion edits | Prioritised Quality Indicator |
|---|---|---|---|---|
| **Respiratory Tract Infection (cont.)** | H19 | Proportion of patients presenting with suspected bacterial CAP given the duration in days of antibiotics recommended in the WHO AWaRe antibiotic book. | Prioritised Supporting Indicator – positive indicator, WHO recommended antibiotic duration | *Proportion of patients presenting with suspected bacterial CAP given the duration in days of antibiotics recommended in the WHO AWaRe antibiotic book.* |
| | H20 | Proportion of patients presenting with suspected bacterial CAP prescribed the total daily dose of IV antibiotics in the WHO AWaRe antibiotic book. | Prioritised Supporting Indicator – positive indicator, WHO recommended antibiotic dose | *Proportion of patients presenting with suspected bacterial CAP prescribed the total daily dose of IV antibiotics in the WHO AWaRe antibiotic book.* |
| | H21 | Proportion of patients with all bacterial CAP given IV/oral Watch antibiotics. | Not Prioritised – duplicate measurement of H16 | |
| **Diarrhoea** | H22 | Proportion of patients with severe acute bloody diarrhoea given the appropriate antibiotic drug choice ~~and duration~~ according to the WHO AWaRe antibiotic book. | During Tidying Process – Remove duration from the indicator as it is measured in H23<br>Prioritised – positive indicator, WHO recommended antibiotic choice | Proportion of patients with severe acute bloody diarrhoea given the appropriate antibiotic choice according to the WHO AWaRe antibiotic book. |
| | H23 | Proportion of patients presenting with severe* acute bloody diarrhoea given the duration in days of IV antibiotics recommended in the WHO AWaRe antibiotic book. | Prioritised Supporting Indicator – positive indicator, WHO recommended antibiotic duration | *Proportion of patients presenting with severe* acute bloody diarrhoea given the duration in days of IV antibiotics recommended in the WHO AWaRe antibiotic book.* |
| | H24 | Proportion of patients presenting with severe* acute bloody diarrhoea prescribed the total daily dose of IV antibiotics in the WHO AWaRe antibiotic book. | Prioritised Supporting Indicator – positive indicator, WHO recommended antibiotic dose | Proportion of patients presenting with severe* acute bloody diarrhoea prescribed the total daily dose of IV antibiotics in the WHO AWaRe antibiotic book. |
| | H25 | Proportion of patients presenting with severe* acute bloody diarrhoea given Access antibiotics. | Not Prioritised – negative indicator, does not align with WHO AWaRe book recommendations for this condition | |

| Hospital Indication | Indicator Number | Original Quality Indicator | Discussion edits | Prioritised Quality Indicator |
|---|---|---|---|---|
| **Diarrhoea (cont.)** | H26 | Proportion of patients presenting with severe* acute bloody diarrhoea given Watch antibiotics. | Not Prioritised – duplicate measurement of H22 | |
| **Intra-abdominal Infections** | H27 | Proportion of patients with mild-moderate intra-abdominal infections (IAIs) given the appropriate IV/oral antibiotic (drug choice and duration) according to the WHO AWaRe antibiotic book. | Not Prioritised – WHO recommendations differ too much between different IAIs to make a meaningful indicator that generalises all IAIs into one category | |
| | H28 | Proportion of patients with mild-moderate intra-abdominal infections (IAIs) given the appropriate IV/oral antibiotic drug according to the WHO AWaRe antibiotic book. | Not Prioritised – WHO recommendations differ too much between different IAIs to make a meaningful indicator that generalises all IAIs into one category | |
| | H29 | Proportion of patients with mild-moderate intra-abdominal infections (IAIs) given the appropriate IV/oral antibiotic duration in days according to the WHO AWaRe antibiotic book. | Not Prioritised – WHO recommendations differ too much between different IAIs to make a meaningful indicator that generalises all IAIs into one category | |
| | H30 | Proportion of patients presenting with intra-abdominal infections (IAIs) (i.e. cholecystitis, cholangitis, acute appendicitis, acute diverticulitis) given IV antibiotics. | Not Prioritised – WHO recommendations differ too much between different IAIs to make a meaningful indicator that generalises all IAIs into one category | |
| | H31 | Proportion of patients presenting with severe* intra-abdominal infections (IAIs) (i.e. cholecystitis, cholangitis, acute appendicitis, acute diverticulitis) given IV Access antibiotic. | Not Prioritised – WHO recommendations differ too much between different IAIs to make a meaningful indicator that generalises all IAIs into one category | |

| Hospital Indication | Indicator Number | Original Quality Indicator | Discussion edits | Prioritised Quality Indicator |
|---|---|---|---|---|
| **Intra-abdominal Infections (cont.)** | H32 | Proportion of patients presenting with severe* intra-abdominal infections (IAIs) (e.g. cholecystitis, cholangitis, acute appendicitis, acute diverticulitis) given IV Watch antibiotic. | Not Prioritised – WHO recommendations differ too much between different IAIs to make a meaningful indicator that generalises all IAIs into one category | |
| | H33 | Proportion of patients with severe IAIs given the appropriate IV/oral antibiotic (drug choice and duration) according to the WHO AWaRe antibiotic book. | Not Prioritised – WHO recommendations differ too much between different IAIs to make a meaningful indicator that generalises all IAIs into one category | |
| | H34 | Proportion of patients with severe IAIs given the appropriate IV/oral antibiotic according to the WHO AWaRe antibiotic book. | Not Prioritised – WHO recommendations differ too much between different IAIs to make a meaningful indicator that generalises all IAIs into one category | |
| | H35 | Proportion of patients with severe IAIs given the appropriate IV/oral antibiotic duration in days according to the WHO AWaRe antibiotic book. | Not Prioritised – WHO recommendations differ too much between different IAIs to make a meaningful indicator that generalises all IAIs into one category | |
| | H36 | Proportion of patients presenting with intra-abdominal infections (IAIs) (i.e. cholecystitis, cholangitis, acute appendicitis, acute diverticulitis) given the duration in days of IV antibiotics recommended in the WHO AWaRe antibiotic book. | Not Prioritised – WHO recommendations differ too much between different IAIs to make a meaningful indicator that generalises all IAIs into one category | |

| Hospital Indication | Indicator Number | Original Quality Indicator | Discussion edits | Prioritised Quality Indicator |
|---|---|---|---|---|
| **Intra-abdominal Infections (cont.)** | H37 | Proportion of patients presenting with intra-abdominal infections (IAIs) (i.e. cholecystitis, cholangitis, acute appendicitis, acute diverticulitis) prescribed the total daily dose of IV antibiotics in the WHO AWaRe antibiotic book. | Not Prioritised – WHO recommendations differ too much between different IAIs to make a meaningful indicator that generalises all IAIs into one category | |
| | H38 | Proportion of patients with all IAIs given IV/oral Watch antibiotics. | Not Prioritised – WHO recommendations differ too much between different IAIs to make a meaningful indicator that generalises all IAIs into one category | |
| **Upper Urinary Tract Infection** | H39 | Proportion of patients with mild* upper urinary tract infection (UTIs) given the appropriate oral antibiotic (drug choice and duration) according to the WHO AWaRe antibiotic book. | Not Prioritised – duplicate measurement of H40 & H41 | |
| | H40 | Proportion of patients with mild* upper urinary tract infection (UTIs) given the appropriate oral antibiotic according to the WHO AWaRe antibiotic book. | Prioritised – positive indicator, WHO recommended antibiotic choice | Proportion of patients with mild* upper urinary tract infection (UTIs) given the appropriate oral antibiotic according to the WHO AWaRe antibiotic book. |
| | H41 | Proportion of patients with mild* upper urinary tract infection (UTIs) given the appropriate oral antibiotic duration in days according to the WHO AWaRe antibiotic book. | Prioritised Supporting Indicator – positive indicator, WHO recommended antibiotic duration | *Proportion of patients with mild* upper urinary tract infection (UTIs) given the appropriate oral antibiotic duration according to the WHO AWaRe antibiotic book.* |
| | H42 | Proportion of patients with severe* upper UTIs given the appropriate IV antibiotics (drug choice and duration) according to the WHO AWaRe antibiotic book. | Not Prioritised – duplicate measurement of H43 & H44 | |

| Hospital Indication | Indicator Number | Original Quality Indicator | Discussion edits | Prioritised Quality Indicator |
|---|---|---|---|---|
| **Upper Urinary Tract Infections (cont.)** | H43 | Proportion of patients with severe* upper UTIs given the appropriate IV antibiotics according to the WHO AWaRe antibiotic book. | Prioritised – positive indicator, WHO recommended antibiotic choice | Proportion of patients with severe* upper UTIs given the appropriate IV antibiotics according to the WHO AWaRe antibiotic book. |
| | H44 | Proportion of patients with severe* upper UTIs given the appropriate IV antibiotics duration in days according to the WHO AWaRe antibiotic book. | Prioritised Supporting Indicator – positive indicator, WHO recommended antibiotic duration | *Proportion of patients with severe* upper UTIs given the appropriate IV antibiotics duration according to the WHO AWaRe antibiotic book.* |
| | H45 | Proportion of patients presenting with upper UTIs given the duration in days of IV antibiotics recommended in the WHO AWaRe antibiotic book. | Not Prioritised – duplicate measurement of H44 | |
| | H46 | Proportion of patients presenting with upper UTIs prescribed the total daily dose of IV antibiotics in the WHO AWaRe antibiotic book. | During Tidying Process – split into two indicators based on severity level<br>Prioritised Supporting Indicator – positive indicator, WHO recommended antibiotic dose | *Proportion of patients presenting with mild/moderate* upper UTIs prescribed the total daily dose of IV antibiotics in the WHO AWaRe antibiotic book.*<br>*Proportion of patients presenting with severe* upper UTIs prescribed the total daily dose of IV antibiotics in the WHO AWaRe antibiotic book.* |
| | H47 | Proportion of patients with all upper UTIs given IV/oral Watch antibiotics. | Not Prioritised – duplicate measurement of H40 & H43 | |
| | H48 | Proportion of patients presenting with upper urinary tract infection (UTI) given IV antibiotics. | Not Prioritised – WHO recommendations between mild/moderate and severe cases for this indication | |
| | H49 | Proportion of patients presenting with severe* upper urinary tract infection (UTI) given IV Access antibiotic. | Not Prioritised – negative indicator, does not align with WHO AWaRe book recommendations for this condition | |
| | H50 | Proportion of patients presenting with severe* upper urinary tract infection (UTI) given IV Watch antibiotic. | Not Prioritised – duplicate measurement of H40 & H43 | |

| Hospital Indication | Indicator Number | Original Quality Indicator | Discussion edits | Prioritised Quality Indicator |
|---|---|---|---|---|
| **Skin/Soft Tissue and Osteoarticular Infection** | H51 | Proportion of patients presenting with complicated skin and soft tissue infections (cSSTIs) (necrotizing fasciitis, pyomyositis) given the appropriate IV antibiotics according to the WHO AWaRe antibiotic book. | Not Prioritised – rare/low burden of necrotizing fasciitis and pyomyositis[6,7] | |
| | H52 | Proportion of patients presenting with complicated skin and soft tissue infections (cSSTIs) (necrotizing fasciitis, pyomyositis) given the duration in days of IV antibiotics recommended in the WHO AWaRe antibiotic book. | Not Prioritised – rare/low burden of necrotizing fasciitis and pyomyositis[6,7] | |
| | H53 | Proportion of patients presenting with complicated skin and soft tissue infections (cSSTIs) (necrotizing fasciitis, pyomyositis) prescribed the total daily dose of IV antibiotics in the WHO AWaRe antibiotic book. | Not Prioritised – rare/low burden of necrotizing fasciitis and pyomyositis[6,7] | |
| | H54 | Proportion of patients presenting with osteoarticular infections (acute bacterial osteomyelitis, septic arthritis) given IV Access antibiotic. | Not Prioritised – rare/low burden of acute bacterial osteomyelitis and septic arthritis based[8,9] | |
| | H55 | Proportion of patients presenting with osteoarticular infections (acute bacterial osteomyelitis, septic arthritis) given IV Watch antibiotic. | Not Prioritised – rare/low burden of acute bacterial osteomyelitis and septic arthritis based[8,9] | |
| | H56 | Proportion of patients presenting with osteoarticular infections (acute bacterial osteomyelitis, septic arthritis) given the duration in days of IV antibiotics recommended in the WHO AWaRe antibiotic book. | Not Prioritised – rare/low burden of acute bacterial osteomyelitis and septic arthritis based[8,9] | |

| Hospital Indication | Indicator Number | Original Quality Indicator | Discussion edits | Prioritised Quality Indicator |
|---|---|---|---|---|
| **Skin/Soft Tissue and Osteoarticular Infection (cont.)** | H57 | Proportion of patients presenting with osteoarticular infections (acute bacterial osteomyelitis, septic arthritis) prescribed the total daily dose of IV antibiotics in the WHO AWaRe antibiotic book. | Not Prioritised – rare/low burden of acute bacterial osteomyelitis and septic arthritis based[8,9] | |
| **Surgical Prophylaxis** | H58 | Proportion of patients who received the appropriate IV surgical prophylaxis according to the WHO AWaRe antibiotic book for bowel surgery/clean or clean-contaminated procedure/contaminated procedure/urologic procedure. | Prioritised – positive indicator, WHO recommended antibiotic choice | Proportion of patients who received the appropriate IV surgical prophylaxis according to the WHO AWaRe antibiotic book for bowel surgery/clean or clean-contaminated procedure/contaminated procedure/urologic procedure. |
| | H59 | Proportion of patients given the duration in days of IV antibiotics recommended in the WHO AWaRe antibiotic book for bowel surgery/clean or clean-contaminated procedure/contaminated procedure/urologic procedure. | Prioritised – positive indicator, WHO recommended antibiotic duration for surgical prophylaxis | Proportion of patients given the duration in days of IV antibiotics recommended in the WHO AWaRe antibiotic book for bowel surgery/clean or clean-contaminated procedure/contaminated procedure/urologic procedure. |
| | H60 | Proportion of patients who received IV surgical prophylaxis at the appropriate timing according to the WHO AWaRe antibiotic book for bowel surgery/clean or clean-contaminated procedure/contaminated procedure/urologic procedure. | Prioritised Supporting Indicator – positive indicator, WHO recommended antibiotic timing for surgical prophylaxis | *Proportion of patients who received IV surgical prophylaxis at the appropriate timing according to the WHO AWaRe antibiotic book for bowel surgery/clean or clean-contaminated procedure/contaminated procedure/urologic procedure.* |
| | H61 | Proportion of patients who received IV surgical prophylaxis at the appropriate timing according to the WHO AWaRe antibiotic book. | Not Prioritised – duplicate measurement of H60 | |

*Red highlighted words reflect changes made during tidying review process
*Italicised priority indicators are supporting indicators measuring duration and dose of antibiotic

Table S11: General Quality Indicators Prioritisation Process: Indicators reviewed and selected as priority and supporting indicators.

| General indication | Indicator Number | Original Quality Indicator | Discussion edits | Prioritised Quality Indicator |
|---|---|---|---|---|
| Patient Level | G1 | The empiric treatment plan should be documented in the medical records and should include clinical diagnosis and indication for antibiotic treatment, dose and route of administration, intended duration of treatment, stop or review date, and IV to oral switch. | Not Prioritised– unclear wording, phrased as guidance rather than a quality indicator | |
| | G2 | Allergy status of the patient including timing, nature and severity of previous exposure/possible allergic reactions to antibiotics and the name/s of the antibiotic should be documented in the medical records. | Not Prioritised– unclear wording, phrased as guidance rather than a quality indicator | |
| | G3 | Antibiotics started for empiric therapy should be reviewed when any microbiological results from the patient become available and the decision to stop or change or continue the empirical therapy should be documented in the medical records. | Not Prioritised– unclear wording, phrased as guidance rather than a quality indicator | |
| | G4 | Antibiotics started for empiric therapy should be reviewed according to clinical response 48-72 hours after the start and the decision to stop or change or continue the empirical therapy should be documented in the medical records. | Not Prioritised– unclear wording, phrased as guidance rather than a quality indicator | |

| | | | | |
|---|---|---|---|---|
| **Patient Level (cont.)** | G5 | Proportion of patients who are eligible for IV to oral switch, in whom intravenous route of administration is switched to the oral route at an appropriate time. | Not Prioritised– unclear wording | |
| | G6 | Any toxicity/adverse reaction to an antibiotic (including type, duration of symptoms, time of onset since antibiotic administration) should be documented in medical records. | Not Prioritised– unclear wording, phrased as guidance rather than a quality indicator | |
| | G7 | Blood cultures should be performed in all patients with severe bacterial infections (sepsis or septic shock, severe pneumonia, meningitis, severe IAIs, severe UTIs) ideally before starting antibiotics or as soon as possible if not sent before. | Not Prioritised– unclear wording, phrased as guidance rather than a quality indicator | |
| | G8 | Proportion of patients with suspected multi-drug-resistant Gram-negative infections given Reserve antibiotics for empiric use according to the WHO AWaRe antibiotic book. | Prioritised Supporting Indicator – important measurement of suspected multi-drug-resistant Gram-negative infections | *Proportion of patients with suspected multi-drug-resistant Gram-negative infections given Reserve antibiotics for empiric use according to the WHO AWaRe antibiotic book.* |
| | G9 | Proportion of patients with suspected multi-drug-resistant infections given Reserve antibiotics for empiric use according to the WHO AWaRe antibiotic book. | Not Prioritised– duplicate measurement of G8 & G11 | |
| | G10 | Proportion of patients with severe illness given Reserve antibiotics for empiric use according to the WHO AWaRe antibiotic book. | Not Prioritised– unclear wording, Reserve antibiotic use for “severe illness” is not in line with WHO recommendations | |

| | | | | |
|---|---|---|---|---|
| **Patient Level (cont.)** | G11 | Proportion of patients with suspected multi-drug-resistant Gram-positive infections given Reserve antibiotics for empiric use according to the WHO AWaRe antibiotic book. | Not Prioritised – negative indicator, does not align with WHO AWaRe book recommendations for this condition | |
| **Population Level** | G12 | Use of oral Access and Watch antibiotics (split by AWaRe group) measured in DID (Defined daily doses per 1000 inhabitants per day) in Primary Care. | Prioritised Supporting Indicator – important measurement of Access and Watch use in primary care | *Use of oral Access and Watch antibiotics (split by AWaRe group) measured in DID (Defined daily doses per 1000 inhabitants per day) in Primary Care.* |
| | G13 | Use of IV and oral Access and Watch and Reserve antibiotics (split by AWaRe group) measured in DDD (Defined daily doses) per 100 hospital admissions per day. | Not Prioritised– duplicate measurement of G14 | |
| | G14 | Use of IV and oral Access and Watch and Reserve antibiotics (split by AWaRe group) measured in DDD (Defined daily doses) per 100 patient days (hospital occupied bed days). | Prioritised Supporting Indicator – important measurement of Access, Watch and Reserve use in hospitals | *Use of IV and oral Access and Watch and Reserve antibiotics (split by AWaRe group) measured in DDD (Defined daily doses) per 100 patient days (hospital occupied bed days).* |
| | G15 | At least 80% of total oral antibiotic use in primary care should be Access antibiotics. | Not Prioritised – negative indicator, does not align with WHO AWaRe book recommendations for this condition | |
| | G16 | Percentage of total oral Access antibiotic use. | Prioritised – important measurement of Access antibiotic use | |
| | G17 | At least 70% of total country level antibiotic consumption should be Access antibiotics. | Not Prioritised – Global target not quality indicator | |

| | | | | |
|---|---|---|---|---|
| **Population Level (cont.)** | G18 | Proportion of antibiotics given for respiratory infections during the high-prevalence season (e.g. winter-summer or wet-dry)<br>compared to<br>The proportion of antibiotics given for respiratory infections given during the low-prevalence season (e.g. summer-winter or dry-wet) | Not Prioritised – Difficult to align with AWaRe book guidance as many RTIS do not require antibiotic | |
| | G19 | Percentage of total country level Access antibiotic use | Prioritised – WHO recommendation that can easily be adapted locally | Percentage of total country level Access antibiotic use |
| | G20 | Ratio of oral amoxicillin measured in DID (Defined daily doses per 1000 inhabitants per day) in primary care to all oral antibiotics in primary care measured in DID excluding amoxicillin (including amoxicillin-clavulanic acid) | Not Prioritised – Difficult to account for differences in case mix in terms of target setting | |
| | G21 | Ratio of oral “Not Recommended” antibiotic measured in DID (Defined daily doses per 1000 inhabitants per day) to all oral antibiotics measured in DID (J01) | Not Prioritised – negative indicator, does not align with WHO AWaRe book recommendations | |
| | G22 | Number of primary care prescriptions >5 days for amoxicillin (J01CA04)<br>Over<br>Total number of primary care antibiotic prescriptions for amoxicillin<br>Proportion of amoxicillin prescriptions >5 days | During Tidying Process: reword indicator to “Proportion of Amoxicillin prescriptions > 5 days”<br>Not Prioritised – negative indicator, does not align with WHO AWaRe book recommendations | |

*Red highlighted words reflect changes made during tidying review process
*Italicised priority indicators are supporting indicators measuring duration and dose of antibiotic
